# Stratification of hypertensive COVID-19 patients by quantitative NMR spectroscopy of serum metabolites, lipoproteins and inflammation markers

**DOI:** 10.1101/2022.12.20.22283729

**Authors:** Jasmin Kazenwadel, Georgy Berezhnoy, Claire Cannet, Hartmut Schäfer, Tobias Geisler, Anne-Katrin Rohlfing, Meinrad Gawaz, Uta Merle, Christoph Trautwein

## Abstract

**Background:** The exact pathophysiology of humans suffering from the multifaceted SARS-CoV-2 infection is not yet conclusively understood and risk stratification is needed. Novel diagnostic approaches like the nuclear magnetic resonance spectroscopy (NMR) based quantification of metabolites, lipoproteins, and inflammation markers has helped to identify typical alterations in the blood serum of COVID-19 patients. However, important confounders such as age, sex, and comorbidities, which strongly influence the metabolome, were often not considered. Therefore, the aim of this NMR study was to consider gender, as well as arterial hypertension (AHT) which affects more than 1.2 billion people worldwide, when investigating COVID-19-positive serum samples in a large age-matched cohort. As AHT is a risk factor for severe COVID-19 disease, this study focuses on comparing metabolomic characteristics of COVID-19 patients with and without AHT.

**Methods and Findings:** NMR serum data from 329 COVID-19 patients were compared with 305 individuals from a healthy age and sex-matched control cohort. 134 of the 329 COVID-19 patients were affected by AHT. These were analyzed together with NMR data from 58 hypertensives without COVID-19. In addition to metabolite, lipoprotein, and glycoprotein data from NMR, common laboratory parameters were considered. Statistical comparison of the COVID-19 cohort with the control cohort reproduced results of previous studies. However, several differences emerged when AHT was considered. Especially, the previously described triglyceride-rich lipoprotein profile was no longer observed in COVID-19 patients, nor was an increase in ketone bodies. Typical metabolic changes that were apparent in COVID-19 patients in both sexes and with AHT were an increase in C-reactive protein (CRP) and the ratio of total glycoprotein (Glyc) to supramolecular phospholipids composite (SPC) which is an inflammatory NMR parameter. Further alterations were a decrease in glutamine, leucine, isoleucine, and lysine, citric acid, HDL-4 particles, and total cholesterol. Typical metabolic cardiovascular risk markers could be detected in hypertensive COVID-19 patients, as well as higher inflammatory NMR parameters than in normotensive COVID-19 patients.

**Conclusion:** We could show that a more precise picture of COVID-19 blood serum parameters emerge when AHT is considered which accordingly should be included in future studies and would help for a refined patient stratification.

## Introduction

To date, the SARS-CoV-2 virus, which emerged in Wuhan, China in December 2019, has infected more than 612 million people worldwide and caused 6.5 million deaths from December 2019 to September 2022 (WHO). COVID-19 infection can manifest in a wide range of symptoms with varying disease severity. Patients often report mild flu-like symptoms or asymptomatic courses. In some cases, however, intensive medical treatment may be necessary or the infection may be fatal. Furthermore, various complications can arise during the course of the disease, which can affect multiple organ systems. These include sepsis, acute respiratory distress syndrome (ARDS) (1), and thromboembolic events such as pulmonary emboli (2) or myocardial infarction (3). Since the beginning of the pandemic, there have been extensive efforts to identify causes and risk factors for severe COVID-19 progression. Several studies have demonstrated that higher age, male gender, and pre-existing conditions such as arterial hypertension (AHT) or diabetes mellitus (DM) may be risk factors for a more severe COVID-19 course and hospitalization. (4, 5).

One way to better understand COVID-19 disease is the use of novel quantitative omics approaches such as nuclear magnetic resonance (NMR) spectroscopy. With this method, a variety of metabolites and lipoproteins can be rapidly quantified in serum from numerous samples. Thus, a metabolic fingerprint of a disease can be identified, which can help to better understand pathogenesis, therapeutic effects, and to identify biomarkers for a more comprehensive diagnosis (6, 7). For COVID-19, several studies have investigated the metabolome and lipoproteome of patient sera both in the acute stage of infection and in the so-called post-acute COVID syndrome. In these studies, it is apparent that there are diverse changes in metabolites during COVID-19 infection. These include a decrease in various amino acids such as histidine, glutamine, lysine, methionine and tyrosine, whereas the amino acid phenylalanine showed increased levels in the studies in COVID-19 patients, and furthermore a general increase in ketone bodies was observed (8-10). COVID-19 patients also showed signs of dyslipidemia with increased levels of very low density lipoprotein (VLDL) particles, triglycerides in total plasma and in lipoprotein subgroups, and a decrease in high density (HDL) and low density lipoprotein (LDL) major and subgroups, which include phospholipids, cholesterol, and apolipoproteins A1 and A2 (8, 11-14). Also, the ABA1 ratio (apolipoprotein B100 divided by apolipoprotein A1) which is considered a cardiovascular risk marker (15), was increased in several COVID-19 cohorts (12, 13). In addition to the quantification of metabolites and lipoproteins in COVID-19 patients, it was possible to make statements about the outcome of patients (9). Accordingly, in hospitalized patients with positive outcome, i.e., in Baranovicova et al. survivors without respiratory deterioration or mechanical ventilation, normalization of energy metabolism occurred more rapidly, as well as an increase in glutamine, than in patients with negative outcome (deterioration of respiratory situation with need for respiratory support or death). Furthermore, NMR technology has identified markers that may be indicative of a severe COVID-19 progression (10, 16-18). As such, above all, increased ketone bodies (16), phenylalanine (16, 17) and pronounced dyslipidemia with increased ABA1 ratio, decreased HDL cholesterol (16) and decreased apolipoprotein A1 (18) had been mentioned. These markers could help to predict response to therapy (16, 19). This was investigated in these two studies with the IL-6 inhibitor tocilizumab, where under and after therapy, respectively, an approximation of the metabolic/lipoproteomic parameters of severe COVID-19 cases to the levels of milder cases was observed. Regarding the post-acute COVID syndrome (PACS), which can also present with a wide range of symptoms (20) and precise disease mechanisms are still unclear, NMR spectroscopy has shown that PACS is associated with delayed metabolic phenoreversion and that the metabolome and lipoproteome still differ from healthy controls one to three months after acute infection (21, 22).

In addition to the metabolites and lipoprotein fractions discussed so far, N-acetylated-glycoproteins can be quantified by NMR as well. These include the GlycA signal, which is caused by various acute phase proteins such as haptoglobin, α1-acid glycoprotein, α1-antitrypsin, α1-antichymotrypsin and transferrin. (23). GlycA is combined with GlycB, which represents N-acetylneuraminic acid (24), to form the Glyc signal and thus maps inflammatory processes (25). Furthermore, Glyc is associated with increased cardiovascular risk (26, 27). Elevated Glyc levels were also found in COVID-19 (14). Another recent NMR parameter is the supramolecular phospholipids composite peak (SPC), which correlates mainly with apolipoproteins and HDL subfractions and is thus decreased in COVID-19 sufferers (28).

As mentioned earlier, cardiovascular disease and diabetes mellitus are among the risk factors for more severe COVID-19 progression (4). These diseases also exhibit diverse alterations in the metabolome and lipoproteome. E.g. in patients with essential arterial hypertension, altered amino acid metabolism has been found, with decreased alanine and methionine levels, and increased arginine levels (29). Lactate and pyruvate were also elevated in hypertensives in one study (30). Furthermore, hypertensive patients show increased levels of cholesterol and triglycerides, as well as increased VLDL and LDL particles with reduced HDL and thus also an increased ABA1 ratio. (31). Elevated serum phenylalanine levels and monounsaturated fatty acids were markers of increased cardiovascular risk in another study (32). Changes in amino acid metabolism are also found in type 2 diabetes mellitus (T2DM), indicating impaired energy metabolism (33). In particular, impaired branched chained amino acid metabolism was conspicuous in this study in diabetic patients with late complications.

### Aim of this study

Cardiovascular disease has both a high prevalence and an impact on metabolites and lipoproteins concentrations in blood serum. Previous studies have taken this mostly not into account when investigating the metabolic phenotype of COVID-19 acute disease. Therefore, in this study, NMR data and laboratory parameters from 58 uninfected hypertensive individuals were compared with 216 samples of COVID-19 affected individuals also suffering from hypertension, and the influence of AHT on the previous findings on metabolic characteristics of COVID-19 was investigated. In our analyses, we demonstrated that the stratification for AHT resulted in a well-refined picture of the NMR based metabolic phenotype of COVID-19. Furthermore, 509 samples of COVID-19 affected individuals with and without hypertension were compared to derive conclusions about risks that could drive a severe disease progression. It was found that hypertensive patients with COVID-19 showed metabolic cardiovascular risk factors and increased inflammatory parameters.

## Material and Methods

### Patient cohort & sample collection

The COVID-19 study population (September 7th 2020 to May 17th 2021) consisted of patients monitored with the “Coronataxi digital early warning” (CDEW) system deployed in Rhein-Neckar County and Heidelberg, Germany - an outpatient care system consisting of remote digital monitoring via a mobile application (with symptom questionnaire and daily pulse oximetry), a medical doctor dashboard and medical care delivery to COVID-19 patients in home quarantine when indicated (34). Visits in home quarantine were initiated when, based on the data obtained from the initial survey and all available self-reported data (e.g. self-reported dyspnea), a medical doctor at the university hospital decided whether a visit in home quarantine by a nurse was indicated and scheduled the visit for the following day.

This was a prospective non-interventional study conducted at the University of Heidelberg. Patients were eligible if they for were diagnosed with SARS-CoV-2 infection and if they had consented to study participation. Written informed consent according to the Declaration of Helsinki was obtained for all patients and the local Ethics committee had approved data collection and analysis (reference number: S-324/2020).

Blood sample collection took place during home visits by nurses (the cohort and study therefore was called “Coronataxi”), which were carried out on medical indication e.g. worsening of symptoms. Therefore, the blood samples were not taken a fastened state and there are various numbers of samples per study participant from zero to three. Also, the time intervals between blood collections varied. From a total of 459 participants in the Coronataxi study, a total of 543 samples from 348 COVID-19 patients was received and investigated by NMR. In interest to get the full statistical power, we took chance of using the NMR data of all samples, even there were several specimens of the same patient. This is a common approach in metabolomics studies. Blood was collected in Li-heparin or EDTA-containing tubes. The specimens were then transported in isolated polystyrene boxes and centrifuged for serum preparation in the afternoon in the laboratory of Heidelberg University Hospital, Germany. A frozen aliquot of the supernatant of these samples was transported to Tübingen, Germany on dry ice and stored at -80 degrees Celsius until further processing and NMR analysis

Besides common laboratory parameters (e.g. creatinine, C-reactive protein, white blood cell count and other) extensive metadata were provided by the University hospital Heidelberg, Heidelberg, Germany e.g. information about anthropometrics, age, medication and pre-existing diseases. The metadata is listed in Table 1 and in the supplementary data S1.

**Table 1.**
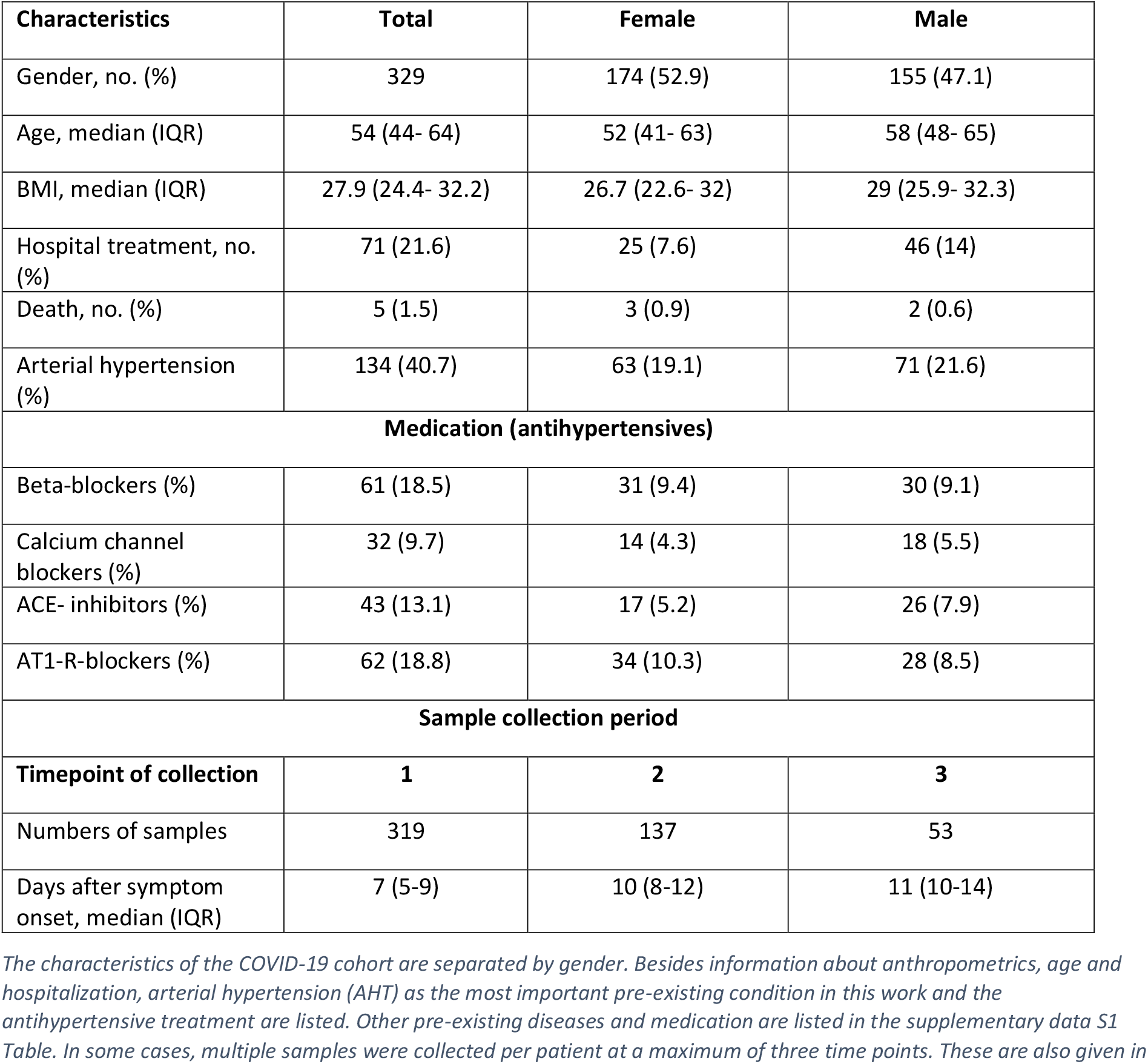
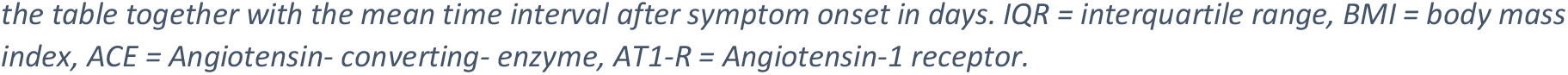
Characteristics of the COVID-19 cohort in total and sample collection.

| Characteristics | Total | Female | Male |
| --- | --- | --- | --- |
| Gender, no. (%) | 329 | 174 (52.9) | 155 (47.1) |
| Age, median (IQR) | 54 (44- 64) | 52 (41- 63) | 58 (48- 65) |
| BMI, median (IQR) | 27.9 (24.4- 32.2) | 26.7 (22.6- 32) | 29 (25.9- 32.3) |
| Hospital treatment, no. (%) | 71 (21.6) | 25 (7.6) | 46 (14) |
| Death, no. (%) | 5 (1.5) | 3 (0.9) | 2 (0.6) |
| Arterial hypertension (%) | 134 (40.7) | 63 (19.1) | 71 (21.6) |
| **Medication (antihypertensives)** |  |  |  |
| Beta-blockers (%) | 61 (18.5) | 31 (9.4) | 30 (9.1) |
| Calcium channel blockers (%) | 32 (9.7) | 14 (4.3) | 18 (5.5) |
| ACE- inhibitors (%) | 43 (13.1) | 17 (5.2) | 26 (7.9) |
| AT1-R-blockers (%) | 62 (18.8) | 34 (10.3) | 28 (8.5) |
| **Sample collection period** |  |  |  |
| **Timepoint of collection** | **1** | **2** | **3** |
| Numbers of samples | 319 | 137 | 53 |
| Days after symptom onset, median (IQR) | 7 (5-9) | 10 (8-12) | 11 (10-14) |

The characteristics of the COVID-19 cohort are separated by gender. Besides information about anthropometrics, age and hospitalization, arterial hypertension (AHT) as the most important pre-existing condition in this work and the antihypertensive treatment are listed. Other pre-existing diseases and medication are listed in the supplementary data S1 Table. In some cases, multiple samples were collected per patient at a maximum of three time points. These are also given in
the table together with the mean time interval after symptom onset in days. IQR = interquartile range, BMI = body mass index, ACE = Angiotensin- converting- enzyme, AT1-R = Angiotensin-1 receptor.

The cohort of the 58 arterial hypertension patients were provided by the interdisciplinary biomaterial- and databank of Würzburg University Hospital, Germany and were age and sex matched with the Heidelberger COVID-19 cohort. They were stored at -80 degrees Celsius, until further processing. The metadata belonging to the serum samples was provided by the institute for clinical epidemiology and biometry Würzburg, Germany. Study enrollment of participants and collection of samples took place between July 2019 and February 2022. The pre-disease AHT was recorded as part of the patient’s medical history. Further metadata contained information about age, sex, medication, diabetes mellitus as co-disease and common laboratory parameters, including renal values (creatinine, blood urea nitrogen, GFR), small blood count, inflammatory values (e.g., CRP), and isolated liver values. The relevant metadata for this work is listed in Table 2. An overview about the antihypertensive treatment is given in Table 3. For the patient recruitment, which took place in Tübingen, Germany, there is an ethics vote with the number 141/2018BO2, dated May 4, 2022. Since the data were kept in Würzburg, Germany, there is a second ethics vote with the number 52/18.

**Table 2.** Characteristics of the COVID-19 cohort, separated by regular blood pressure and AHT, and the AHT control cohort.

| Characteristics | COVID- 19 regular BP | COVID- 19 AHT | AHT control cohort |
| --- | --- | --- | --- |
| Total | 195 | 134 | 58 |
| Gender, no. (%): |  |  |  |
| Female | 111 (56.9) | 63 (47.0) | 29 (50) |
| Male | 84 (43.1) | 71 (53.0) | 29 (50) |
| Age, median (IQR) | 49 (37- 60) | 62 (53- 69) | 57 (52- 61) |
| Hospital treatment, no. (%) | 38 (19.5) | 33 (24.6) | n.a. |
| CRP, median (IQR) | 18.1 (8.7- 48.4) | 21.3 (8.3- 57.8) | 0.4 (0.1- 1.2) |

This table presents the relevant characteristics of the COVID- 19 cohort with regular BP and AHT, as well as the AHT control cohort. Listed are the total patient numbers, gender, age, hospitalization, and the laboratory parameter C-reactive protein (CRP). BP =blood pressure, AHT= arterial hypertension, IQR= interquartile range.

**Table 3:** Antihypertensive treatment of the AHT control cohort.

| Antihypertensives | Total (%) | Female (%) | Male (%) |
| --- | --- | --- | --- |
| Beta-blockers (%) | 42 (72.4) | 21 (36.2) | 21 (36.2) |
| Calcium channel blockers (%) | 22 (37.9) | 11 (19) | 11 (19) |
| ACE- inhibitors (%) | 24 (41.4) | 14 (24.1) | 10 (17.2) |
| AT1-R-blockers (%) | 31 (53.4) | 15 (25.9) | 16 (27.6) |

This table shows the antihypertensive treatment with the four substance classes beta- blockers, calcium channel blockers, ACE- inhibitors, AT1-R-blockers of the 58 patients in the AHT control cohort. ACE = Angiotensin- converting- enzyme, AT1-R = Angiotensin-1 receptor.

For improved statistical analysis, further NMR data (metabolites and lipoproteins) of an age and sex matched healthy control cohort (HC) of 305 subjects were provided by Bruker BioSpin GmbH (Ettlingen, Germany). These data did not contain any clinical laboratory parameters.

### Sample preparation and NMR measurement

The raw NMR spectra were acquired using Bruker’s body fluids NMR methods package [https://www.bruker.com/de/products-and-solutions/mr/nmr-clinical-research-solutions/b-i-methods.html]. The sample preparation was performed following standard operating procedures (SOPs) to perform reproducible results. Briefly, blood serum samples were thawed to room temperature. Then 400 µl of commercially available plasma/serum buffer (Bruker BioSpin GmbH, Ettlingen, Germany) were pipetted into a 1,5 ml sterile Eppendorf tube. Next, an aliquot of 400 µl serum was added to the buffer. After gently shaken, an aliquot of 600 µl of the mixture was transferred into a 5 mm Bruker SampleJet NMR tube. Since only 350 µl was available for the cohort with the 58 hypertensive patients, 320 µl serum aliquot was added to 320 µl Bruker Plasma buffer (resulting in the same 1:1 mixing ratio) and an aliquot of 600 µl was used for measurement.

NMR spectroscopy was performed with a Bruker BioSpin IVDr Avance III HD 600 MHz system. It uses a temperature-controlled autosampler SampleJet™ for gentle handling during the measurements. 1D 1H-NMR spectra of the NOESY experiment (Nuclear Overhauser Spectroscopy) at a temperature of 310 K were acquired. NOESYs were analyzed using Bruker’s software TopSpin (Version 3.6.2) and automatically processed. For the highest standardization and amount of reproducibility the following NMR-based commercial IVDr SOPs (standard operation procedures) were used to collect the data sets for quality control, metabolites, lipoproteins, and inflammatory markers: B.I.BioBankQCTM in Plasma/Serum (Quality Control for Biobank Blood Samples), B.I.Quant-PSTM (quantification of up to 41 metabolites in serum in mmol/L), B.I.LISATM (Lipoprotein analysis for 112 lipoprotein variables in serum), and B.I. PACS™ (quantification of two metabolites, nine lipoprotein variables, SPC and inflammatory markers e.g., GlycA/GlycB and Glyc/SPC ratio which provide information related to various complications in the context of post-acute COVID-19 syndrome).

Lipoprotein concentrations provided by the B.I. LISA™ package are divided in various subfractions broken down by triglycerides (TG), cholesterol (CH), apolipoproteins A1/ -A2, apolipoprotein B100 bearing particles (AB), and total particle numbers (PN). These subfractions exist in the individual lipoprotein classes, very-low-density (VLDL), intermediate density (IDL), low-density (LDL), and high-density lipoproteins (HDL). Additionally, the above mentioned subfractions (TG, CH, A1, A2, AB, PN) are also available in total plasma concentrations (TP). Phospholipid (PL) and free cholesterol (FC) fractions are also provided within the lipoprotein panel. Furthermore, the lipoproteins are subdivided according to size and thus increasing density. VLDL can be subdivided by NMR into five sizes, LDL into six, and HDL into four whereas an increasing number is associated with smaller size or higher density.

Glycoproteins (Glyc A, Glyc B, Glyc) and the supramolecular phospholipid composite (SPC) data from the B.I. PACS™ module were available for the COVID-19 cohort and the AHT control cohort, but not for the HC from Bruker.

### Quality control, Statistics and Data illustration

To increase the quality of the NMR data set, the above-mentioned quality control reports of the individual samples were checked and a total of 26 samples were sorted out based on these, mainly due to a linewidth of > 2.3 Hz. This resulted in a total 509 samples from 329 COVID-19 patients and 58 samples from 58 hypertension patients were investigated.

The software package MetaboAnalyst 5.0 was used for statistical analysis of the obtained quantitative NMR reports. To change the data as little as possible, no normalization or scaling has been applied as NMR measurements are highly reproducible and results are based on absolute quantification. Only log transformation was performed to correct for heteroskedasticity. Features with > 50% missing values have been removed. The other missing values were estimated with k-nearest neighbors (KNN) based on similar features. As univariate test method, volcano plots, which corresponds to a combination of unpaired t-test and fold change (FC) analysis, were used to determine significant alterations of metabolites and lipoproteins. Thereby a p-value threshold of < 0.05 and FC threshold of 1.2 were set. For false discovery rate (FDR) control, Benjamini-Hochberg procedure was used. For multivariate statistics, unsupervised principal component analysis (PCA) was performed to get an overview of the clustering and to identify outliers. Features such as the 2D scores plots, loadings plots, and biplots were used for this purpose. For Table 4, the Biomarker analysis was used to elaborate the most prominent alterations of the respective comparisons. From all significant metabolites, lipoproteins, and laboratory parameters in the volcano plot analyses, those 15 with the highest area under the curve (AUC) were chosen and are listed in Table 4.

**Table 4.**
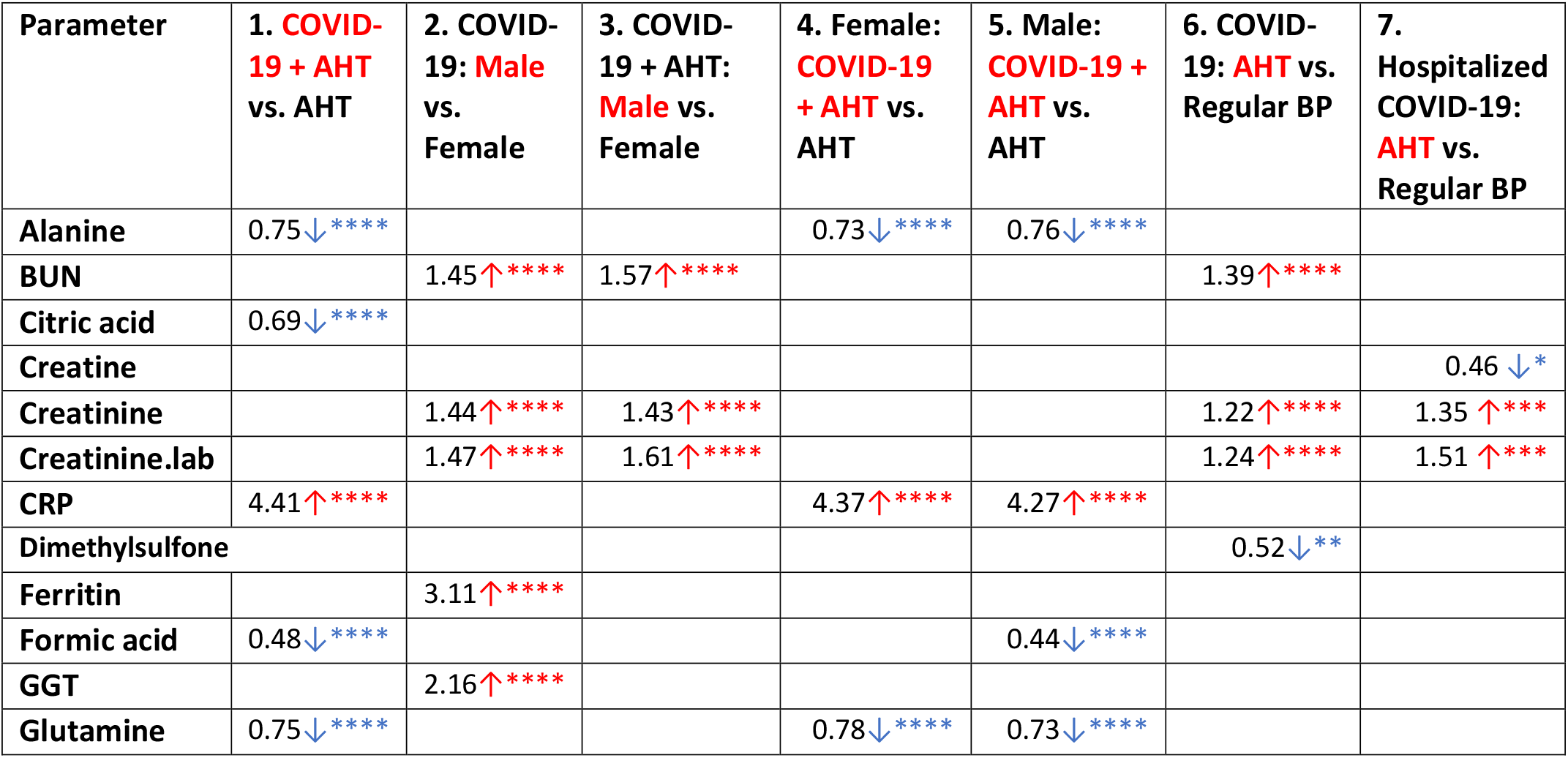
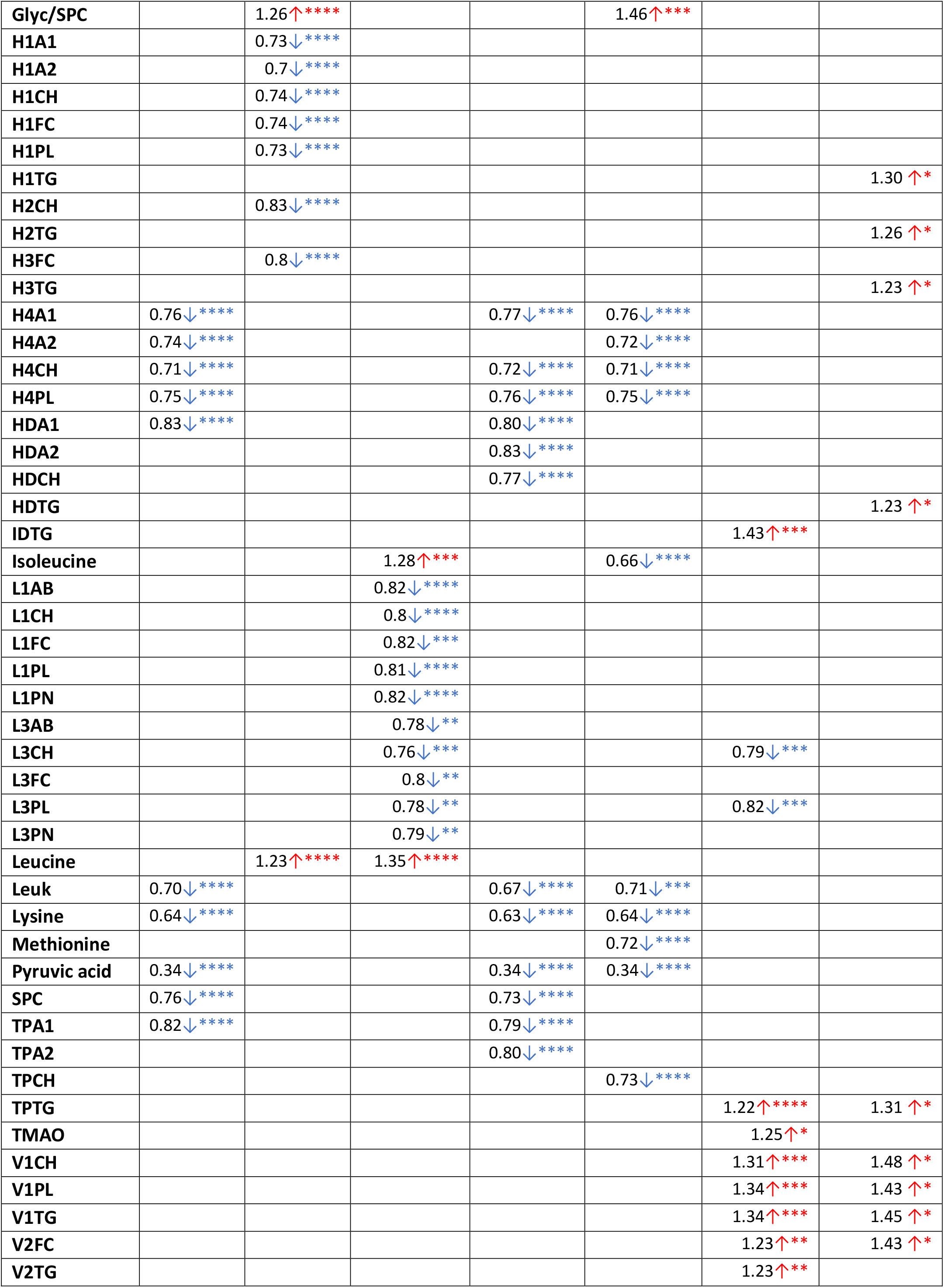
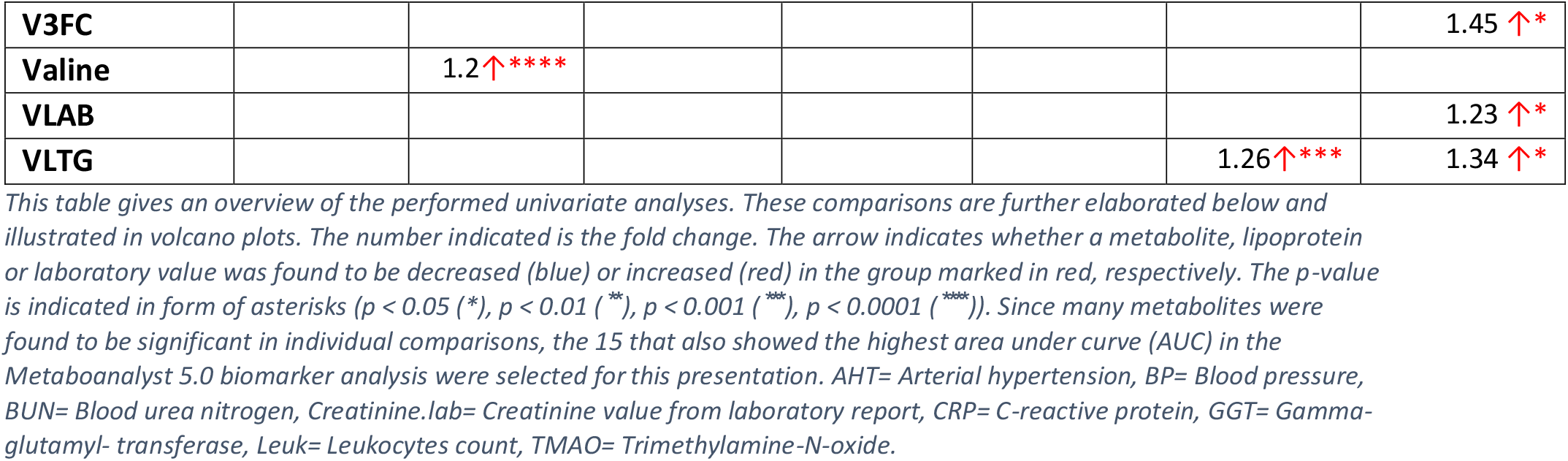
Overview of results of the individual group comparisons in the univariate statistics.

Plots of multivariate statistics were used from MetaboAnalyst 5.0. Further, violin plots and volcano plots included in this article were created with GraphPad Prism 9. The consort diagram (Fig 1) was created with Microsoft PowerPoint 2019.

**Fig 1.**
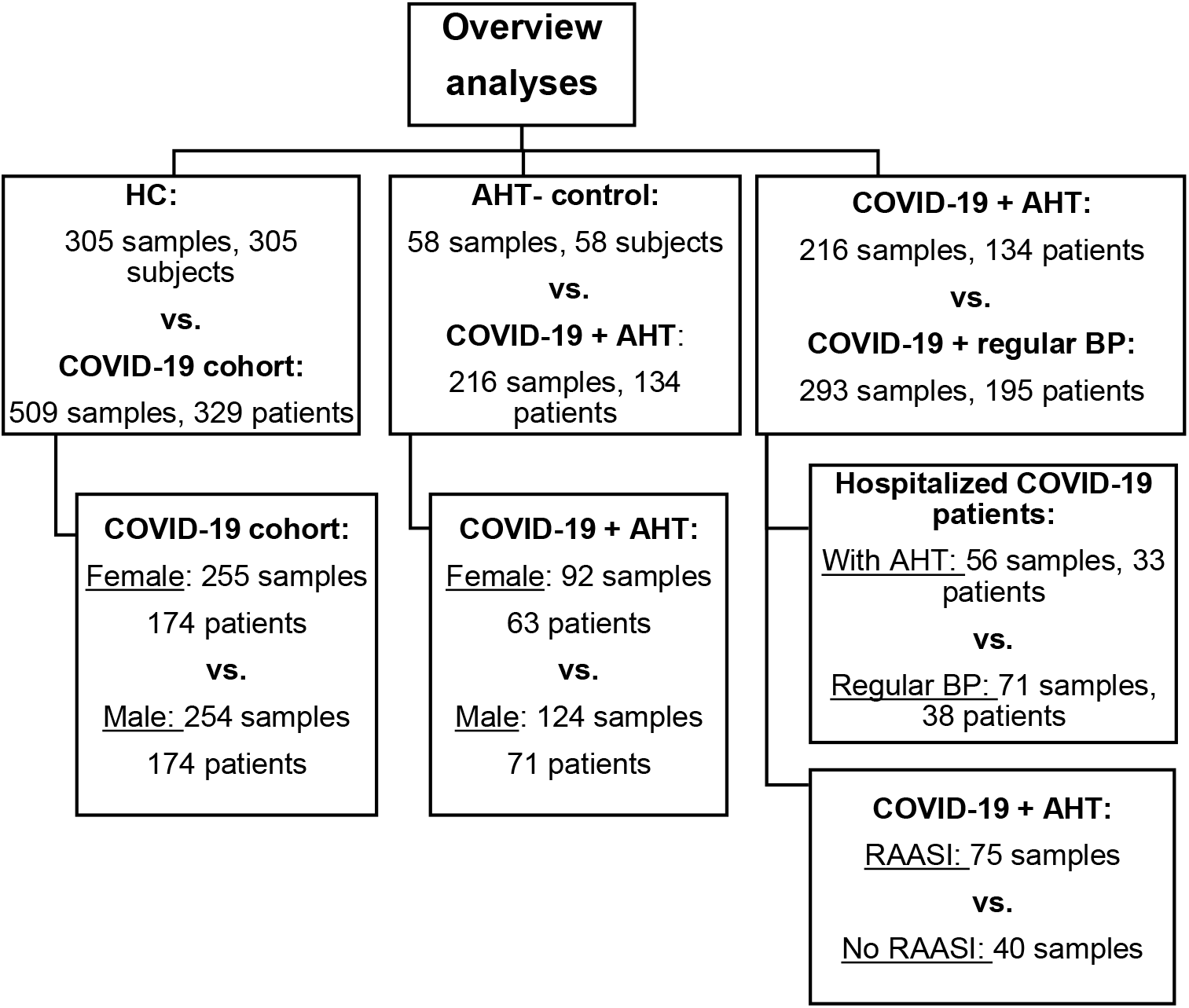
Consort diagram showing an overview of the cohorts. Presented are patient and sample numbers, which were used for the various analyses. HC= Healthy control, AHT= Arterial hypertension, BP= Blood pressure, RAASI= Renin-angiotensin-aldosterone-system inhibitors.

## Results

### Cohort composition

The main characteristics of the Coronataxi cohort are shown in Table 1. As this cohort was divided in a normotensive and hypertensive COVID-19 patient group, some characteristics for these are listed separately in Table 2, along with the AHT control cohort.

### Overview of the performed analyses and results

In this work, NMR data from a healthy cohort (n= 305) were compared with NMR data of the COVID-19 cohort in total (n= 509). Further, NMR data and laboratory parameters of COVID-19 affected individuals with AHT (n=216) were compared with an AHT control cohort (n=58) to investigate the impact of AHT on previous findings on metabolomic, lipoproteomic and inflammatory changes in COVID-19 patients. These comparisons were additionally analyzed separately by gender. NMR data and laboratory parameters were also analyzed from COVID-19 patients with AHT (n=216) and without (n= 293). The same was performed with samples of hospitalized COVID-19 patients, of which 33 individuals (sample n=56) suffered from AHT, while 38 patients (sample n=71) had a regular blood pressure. Last, AHT patients with different antihypertensive treatment were compared. An overview of all performed analyses is given in Fig 1.

Table 4 is intended to provide an overview of the results of the individual univariate analyses. Regardless of gender and prior AHT (columns 1, 4, 5), all COVID-19 patients showed an increase in CRP, decreases in alanine, glutamine, lysine, pyruvate, and in the lipoprotein subgroups H4A1, H4CH, and H4PL. Other common features not included in this presentation, due to a smaller AUC but still showing significant changes in univariate statistics, were an increase in Glyc/SPC ratio, decreased levels of citric acid, isoleucine, and leucine, as well as HDL-subgroups H4A2 and H4FC, total cholesterol (TPCH), total apolipoproteins (TPA1, TPA2), and SPC. Interestingly, COVID-19 groups had decreased leukocyte counts in contrast to the control group. There were differences of laboratory parameters in COVID-19 affected depending on gender (column 2). This included higher levels of creatinine, ferritin, gamma-glutamyl transferase (GGT) and blood urea nitrogen (BUN) in males. It was also noticeable that men had higher inflammatory parameters in the form of Glyc/SPC. The branched chained amino acids (BCAA) leucine and valine were increased in men. HDL subclasses (HDL-1, -2, -3) were decreased. When COVID-19 + AHT affected women and men were compared (column 3), there were some changes. In this subgroup, the BCAAs leucine and isoleucine were increased in male patients, but the changes in lipoproteins now concerned LDL subgroups (mainly the larger particles LDL-1 and -3), which showed lower levels in men than in women. When COVID-19 patients with normal blood pressure and AHT were compared (column 6), metabolic markers associated with AHT were conspicuous in the hypertensive COVID-19 group. These included elevated creatinine and BUN, a triglyceride-rich lipoprotein profile (IDTG, TPTG, VLTG, VLDL-1, -2), and the intestinal metabolite trimethylamine-N-oxide (TMAO). The last mentioned was found to be elevated only in the COVID-19 + AHT group. Hospitalized hypertensive COVID-19 patients (column 7), also showed a triglyceride-rich lipoprotein profile compared with hospitalized normotensive COVID-19 patients. This was characterized by increased total and VLDL triglycerides (TPTG, VLTG), VLDL particles (VLAB, VLDL-1, VLDL-2, VLDL-3), and, in contrast to the whole COVID-19 + AHT group, also by an increase in HDL triglycerides (HDTG, H1TG, H2TG, H3TG). All group comparisons discussed here, as well as additionally the comparison of the whole COVID-19 cohort with Bruker’s HC and the comparison of different antihypertensive treatment groups, will be discussed in detail in the next chapters.

### Differences in the metabolomic phenotypic when comparing total COVID-19 patient cohort with sub-cohort of patients with hypertension

For comparison with previously reported findings of NMR-based alterations of metabolites and lipoproteins, a volcano plot analysis was performed comparing the 305 samples of the HC with the entire Coronataxi COVID-19 cohort, i.e., 509 samples. All metabolic changes named below therefore apply to the comparison of the entire COVID-19 cohort with Bruker’s HC and can be seen in the volcano plot in Fig 2A. In this plot, 60 metabolites and lipoproteins were decreased more than 1.2-fold, whereas 29 were increased. The metabolic changes largely coincided with the literature already cited in the introduction. A triglyceride-rich lipoprotein profile could be detected in the total COVID-19 cohort based on the elevated levels of total plasma triglyceride (TPTG), other triglyceride subgroups (IDTG, L1TG), all major VLDL groups (VLAB, VLPN, VLCH, VLFC, VLTG, and VLPL), and several VLDL subgroups (e.g. V1TG, V2CH, V3TG, V5FC, for example) in all sizes except VLDL-4. On the other hand, a reduction in total plasma apolipoproteins A1 and A2 and all major HDL-groups (HDA1, HDA2, HDCH, HDFC, HDPL), except HDTG, was observed in the COVID-19 cohort. All four sizes were present, with HDL-3 and -4 as the most significant subgroups, of which the apolipoprotein, phospholipid, and cholesterol subgroups were particularly represented. Furthermore, total plasma cholesterol (TPCH), several LDL major (LDCH, LDFC, LDPL), and subgroups, were shown to be decreased, this being mainly true for denser LDL subgroups (LDL-3, -4, -5, -6), as well as for the phospholipid and cholesterol subgroups (e.g., L4CH, L5PL, L6FC). For metabolites, we observed a decrease over 1.2-fold in several amino acids, including glutamine, lysine, histidine, leucine, and isoleucine. Another reduced metabolite was citric acid. In contrast, ketone bodies (3-hydroxybutyrate, acetone), the amino acids phenylalanine and glutamic acid, and the metabolites succinic acid and dimethylglycine were increased in the COVID-19 cohort.

**Fig 2.**
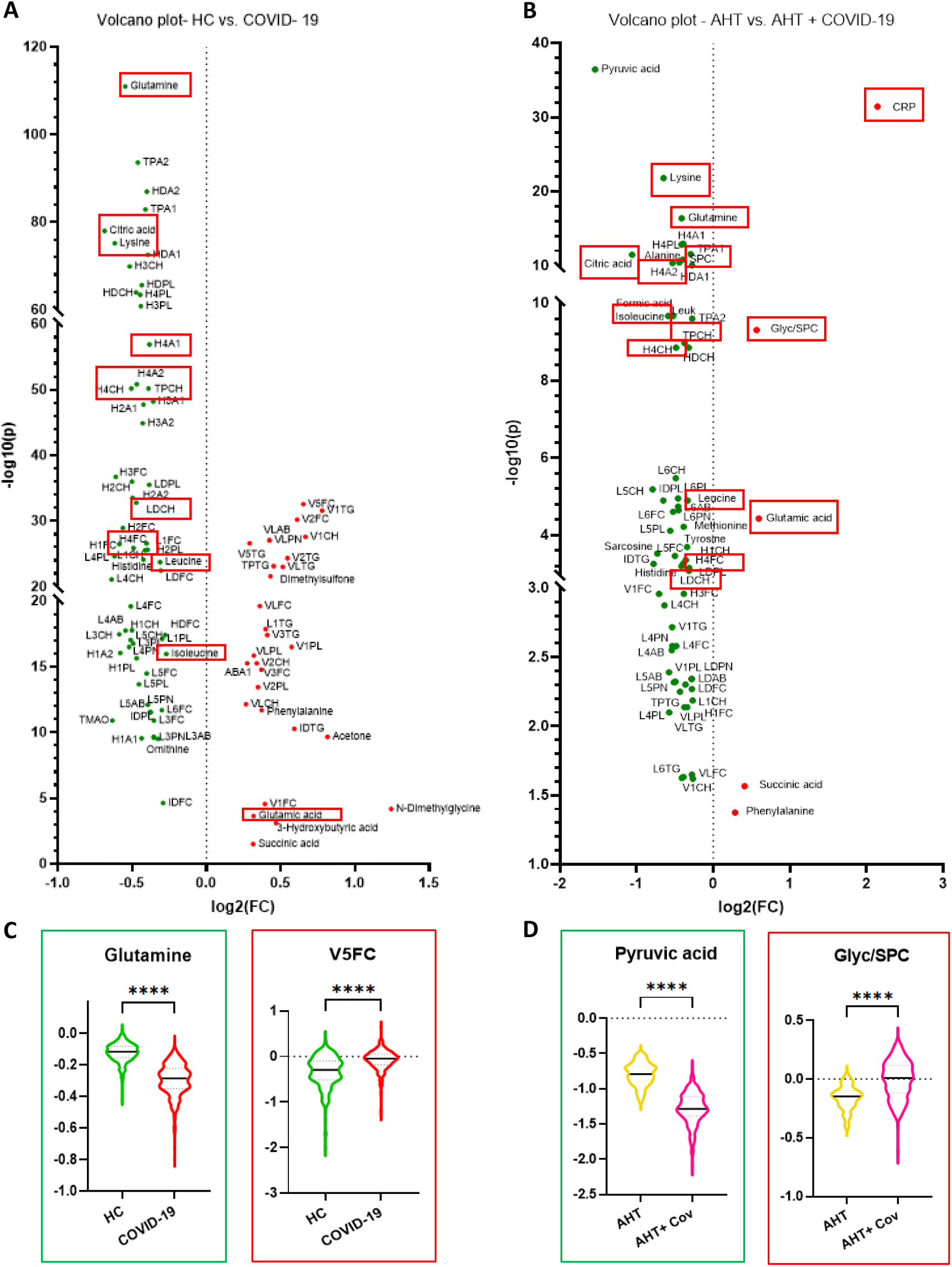
Univariate comparisons by means of volcano plots showing the differences of the metabolome and lipoproteome in COVID-19 + AHT patients compared to the COVID-19 cohort in total. FC > 1.2, p < 0.05, FDR < 0.01: The red features are significantly increased in the COVID-19 cohorts, respectively, while the green ones are significantly decreased in COVID-19 affected. The features in red boxes showed up repeatedly in these and following comparisons and were therefore typical altered in COVID-19 patients. (A) shows the comparison between the COVID-19 cohort in total with the HC. (B) shows the comparison between hypertensive COVID-19 patients and uninfected hypertensive individuals. In (C) and (D) violin plots show the decreased respectively increased NMR parameters with the highest significance for each comparison. The y-axis shows the normalized concentration. HC= Healthy control, AHT= Arterial hypertension, Cov= COVID-19, CRP= C-reactive protein, Leuk= Leukocytes count.

To see whether the results described above could be reproduced when stratifying for AHT, the COVID-19 cohort was divided into a hypertensive and a normotensive sub-cohort. The COVID-19 cohort with AHT was compared with an AHT control group without COVID-19 using another volcano plot (Fig 2B). We could identify 60 metabolites, lipoproteins, and laboratory parameters which showed a decrease of more than 1.2 -fold in the COVID-19 + AHT cohort (as in the previous comparison), while 5 showed an increase (versus 29 increased features in the comparison COVID-19 in total vs. HC). The main differences between those two comparisons were that we could not observe a triglyceride-rich lipoprotein profile in the COVID-19 + AHT cohort. Only the inflammatory parameters CRP and Glyc/SPC ratio, the amino acids glutamic acid and phenylalanine, and the citrate cycle metabolite succinic acid were elevated. The above-described increase in ketone bodies did not show up in the COVID-19 + AHT group. Furthermore, the significant reductions in HDL-fractions in COVID-19 + AHT patients were mainly concentrated on HDL-4 particles (H4A1, H4A2, H4CH, H4FC, H4PL), in contrast to the whole COVID-19 group. On the other hand, there were some features, which these two comparisons had in common, including decreases in amino acids lysine, glutamine, histidine, leucine, and isoleucine in the SARS-CoV-2 infected patients with and without AHT. In the COVID-19 + AHT cohort, additionally, the amino acids alanine, methionine, and tyrosine were decreased. Another common metabolite was citric acid, which showed decreased levels in COVID-19 patients in total and in COVID-19 + AHT. As in the whole COVID-19 cohort, also in the hypertensive group, a reduction of TPCH and several LDL-major-(LDAB, LDCH, LDFC, LDPL) and subgroups could be observed. Again, mostly denser particles (LDL-4, -5, -6) and phospholipid and cholesterol fractions of LDL were present. Interestingly, a few major VLDL groups (VLFC, VLPL, VLTG) and isolated VLDL-1 particles were also reduced in the COVID-19 + AHT cohort.

### Gender differences in COVID-19 patients with and without AHT

In Fig 3A, a volcano plot was used to compare 155 (254 samples) male and 174 (255 samples) female COVID-19 patients from the cohort in total. Overall, 26 features were significantly increased in male COVID-19 patients, whereas 19 were decreased. Some gender-specific laboratory parameters were altered, such as creatinine, blood urea nitrogen (BUN), ferritin, and the liver values GGT and GOT, which showed higher values in men than in women. Furthermore, males with COVID-19 showed higher levels of Glyc/SPC ratio, BCAAs leucine, isoleucine, and valine, ketone bodies (acetones, acetoacetic acid, 3-hydroxybutyric acid), and various VLDL-subgroups, including triglyceride (VLTG, V1TG, V2TG), phospholipid (V1PL, V2PL, V3PL), and cholesterol (V1FC, V1CH, V2FC, V2CH, V3FC) fractions. In comparison, women showed higher values in HDL-subgroups. This was true for apolipoprotein (H1A1, H1A2), cholesterol (H1CH, H1FC, H2CH, H2FC, H3FC), phospholipid (H1PL), and triglyceride fractions (H1TG). HDL-4 did not stand out in the gender comparison. Furthermore, LDL particles were more abundant in female COVID-19 patients. Among them were the larger LDL-subgroups, like LDL-1 (L1CH), LDL-2 (L2CH, L2PL), and LDL-3 (L3AB, L3CH, L3FC, L3PN, L3PL).

**Fig 3.**
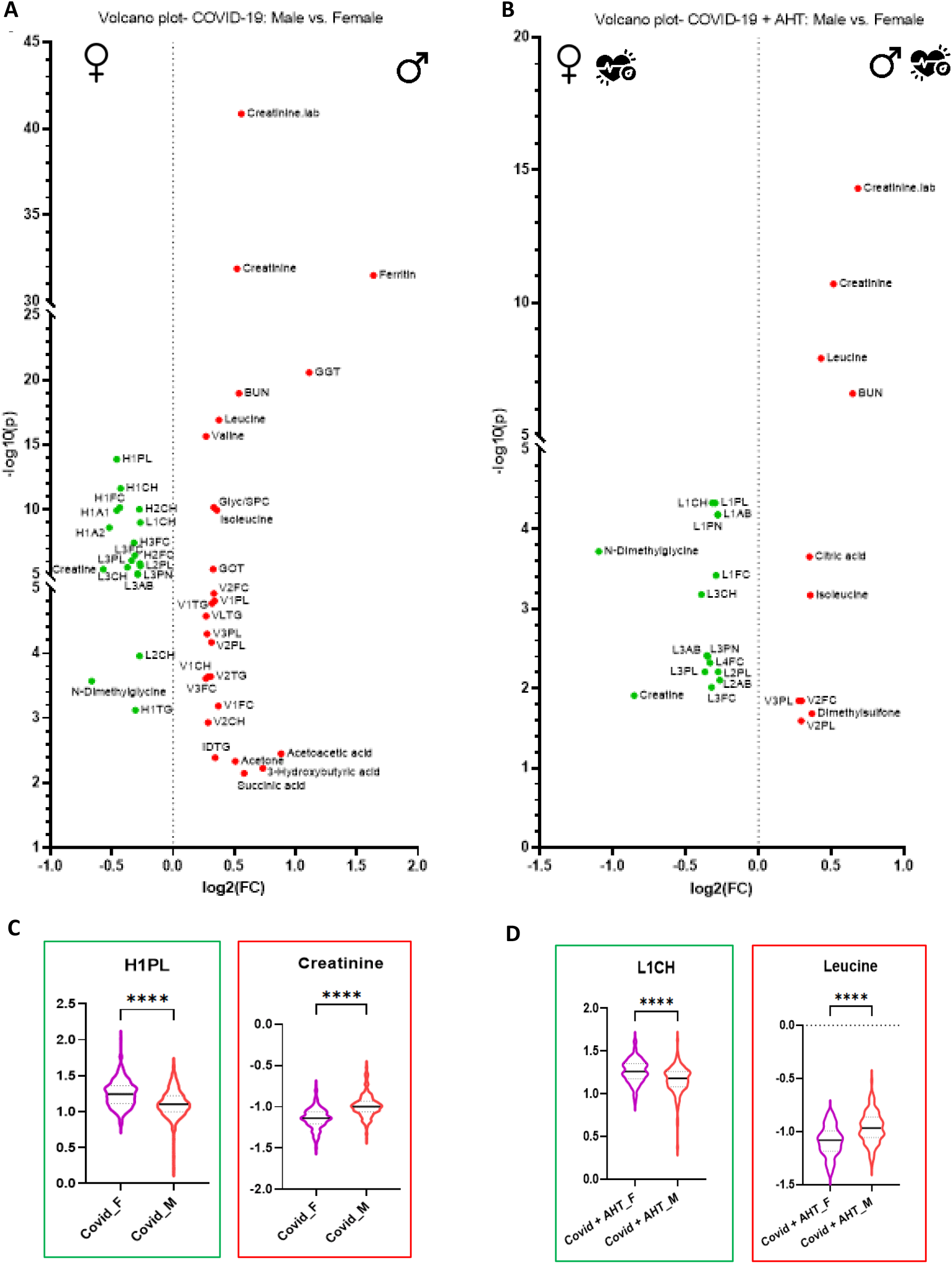
Univariate comparisons by means of volcano plots showing gender differences of COVID-19 patients with and with-out AHT. FC > 1.2, p < 0.05, FDR < 0.01: The red features are significantly increased in men, respectively, while the green ones are significantly decreased in men, compared to women. (A) shows the comparison between the genders in the COVID-19 cohort in total. (B) shows the comparison between the genders in the COVID-19 + hypertension cohort. In (C) violin plots show the decreased respectively increased NMR parameters with the highest significance. In (D) the left violin plot shows L1CH, as the lipoprotein with the highest significance, which is decreased in males. The right violin plot shows leucine, as increased NMR parameter in males, with the second highest significance after creatinine. The y-axis shows the normalized concentration. AHT= Arterial hypertension, BUN= Blood urea nitrogen, Creatinine.lab= Creatinine from laboratory report, GGT= Gamma-glutamyl-transferase, GOT= Glutamate-oxalacetate-transferase, F= Female, M= Male.

To examine the effect of AHT on metabolic sex differences in COVID-19 patients, the COVID-19 + AHT cohort was divided by sex. A volcano plot was performed with data from 124 samples from 71 men and 92 samples from 63 women with COVID-19 + AHT (Fig 3B). This comparison in the AHT subgroup revealed less significant differences than present in the total COVID-19 cohort. 10 features (vs. 26 in the male COVID-19 group in total) were significantly increased in males compared to females; 15 features (vs. 19 in the male COVID-19 cohort in total) were decreased in male patients. The triglyceride-rich lipoprotein profile of men now was less pronounced, only isolated VLDL particles were elevated (V2FC, V2PL, V3PL). Ketone bodies no longer stood out as significant, nor did the Glyc/SPC ratio. Men showed higher levels of the BCAAs leucine and isoleucine, as well as citric acid, and dimethyl sulfone. Women continued to show higher values in the LDL fractions, but not in the HDL particles. Among them, LDL-1 (L1AB, L1CH, L1FC, L1PL, L1PN), LDL-2 (L2AB, L2PL), LDL-3 (L3AB, L3CH, L3FC, L3PL, L3PN), and LDL-4 (L4FC) were present.

The comparisons of COVID-19 + AHT with an AHT control group, as well as the gender comparisons, identified characteristics that are independent of gender and prior AHT disease and thus typical of COVID-19 infection in our cohort. These are shown as outlined in red in Fig 2 and 4. In the volcano plots, male and female hypertensive patients with and without COVID-19 were compared. There were 29 samples from 29 female hypertensive patients without COVID-19 and 92 samples from 63 female hypertensive patients with COVID-19 and AHT. In male patients, 29 samples from 29 hypertensive patients without and 124 samples from 71 patients with COVID-19 and AHT were also compared. In these analyses, metabolites and lipoproteins that were also apparent in the previous univariate statistics stood out. These included an increase in Glyc/SPC, glutamic acid and CRP, and a decrease in amino acids leucine, isoleucine, glutamine, and lysine, citric acid, and HDL-4 fractions (H4A1, H4A2, H4CH, H4FC, H4PL), total cholesterol (TPCH), and SPC.

**Fig 4.**
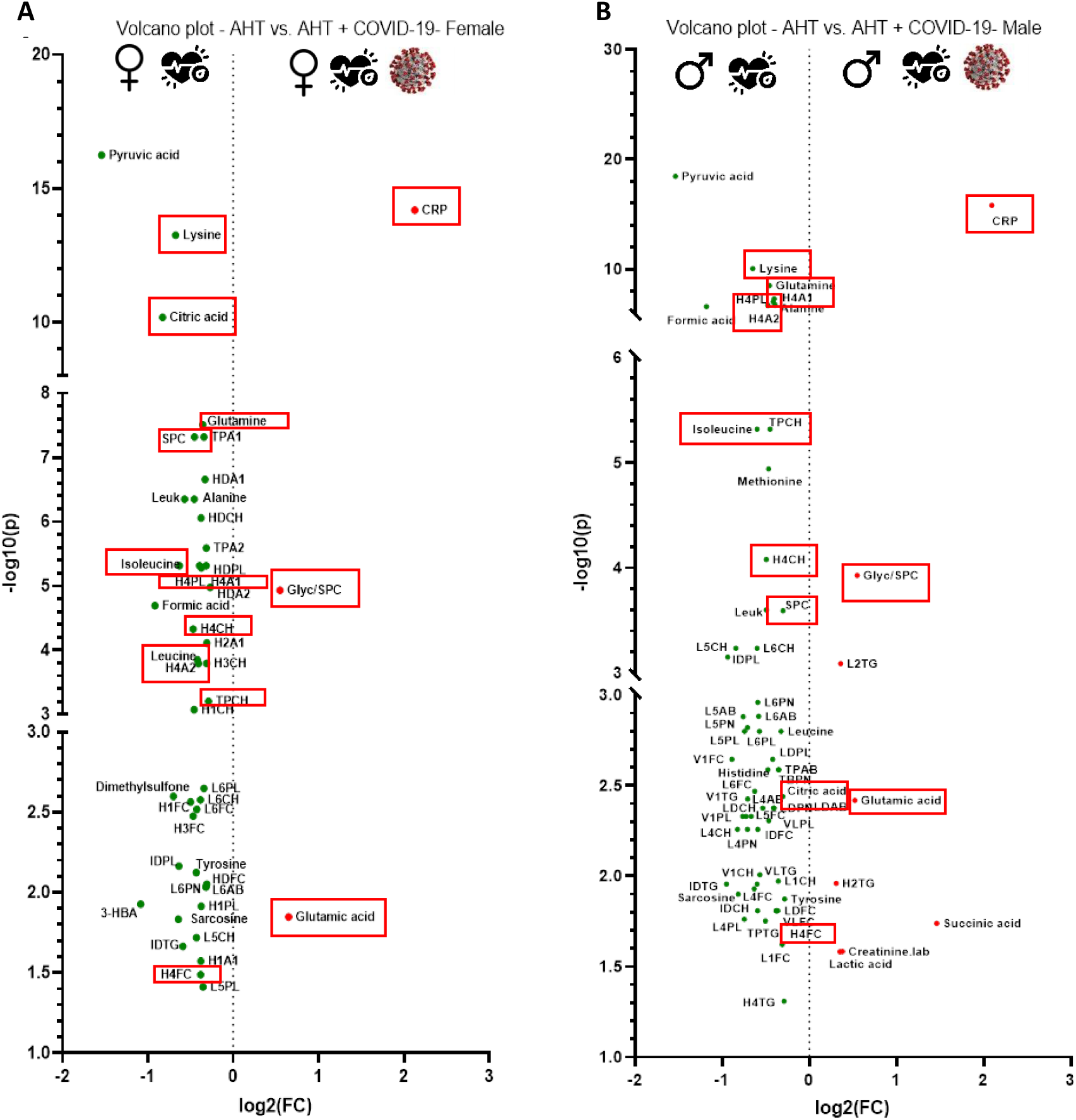
Univariate comparison of AHT cohorts with and without COVID-19, separated by gender, respectively. FC > 1.2, p < 0.05, FDR < 0.01: The red features are significantly increased in COVID-19 patients, while the green ones are decreased. Plot (A) shows the comparison of AHT vs. AHT + COVID-19 for females, plot (B) shows this comparison for male gender. Features in red boxes are common in both genders and in COVID-19 affected considering AHT. AHT= Arterial hypertension, Creatinine.lab= Creatinine from laboratory report, CRP= C-reactive protein, 3-HBA= 3-Hydroxybutyric acid.

### Hypertensive COVID-19 patients show NMR-based metabolomic and lipoproteomic characteristics of an increased cardiovascular risk

The aim of the next analysis was to compare hypertensive and normotensive COVID-19 patients to investigate if there are metabolic alterations between those two groups which could indicate a higher risk for a severe COVID-19 disease course. 216 samples from 134 hypertensive COVID-19 patients were compared with 293 samples from 195 normotensive COVID-19 patients. A volcano plot (Fig 5) was performed, where 15 metabolites, lipoproteins, and laboratory parameters were increased more than 1.2-fold. Two lipoprotein subclasses (L3CH, L3PL) and one metabolite (dimethyl sulfone) were reduced. The highest significant alterations were seen in the elevated renal values in the hypertensive COVID-19 group. BUN and creatinine.lab corresponded to standard laboratory parameters, whereas creatinine was part of the NMR-based B.I. QUANT-PS report. In addition, significant increases in triglyceride-rich VLDL particles were observed, especially in the larger and less dense subgroups (VLDL-1, -2, -3), as well as in the triglyceride subfractions (VLTG, V1TG, V2TG). Total plasma triglyceride (TPTG) and IDL triglyceride fraction (IDTG) also showed elevated levels. Regarding metabolites, the citrate cycle metabolite succinic acid was increased, as in other analysis. The metabolite trimethylamine-N-oxide (TMAO), which is produced by intestinal bacteria and is considered a cardiovascular risk marker, only stood out in this analysis.

**Fig 5.**
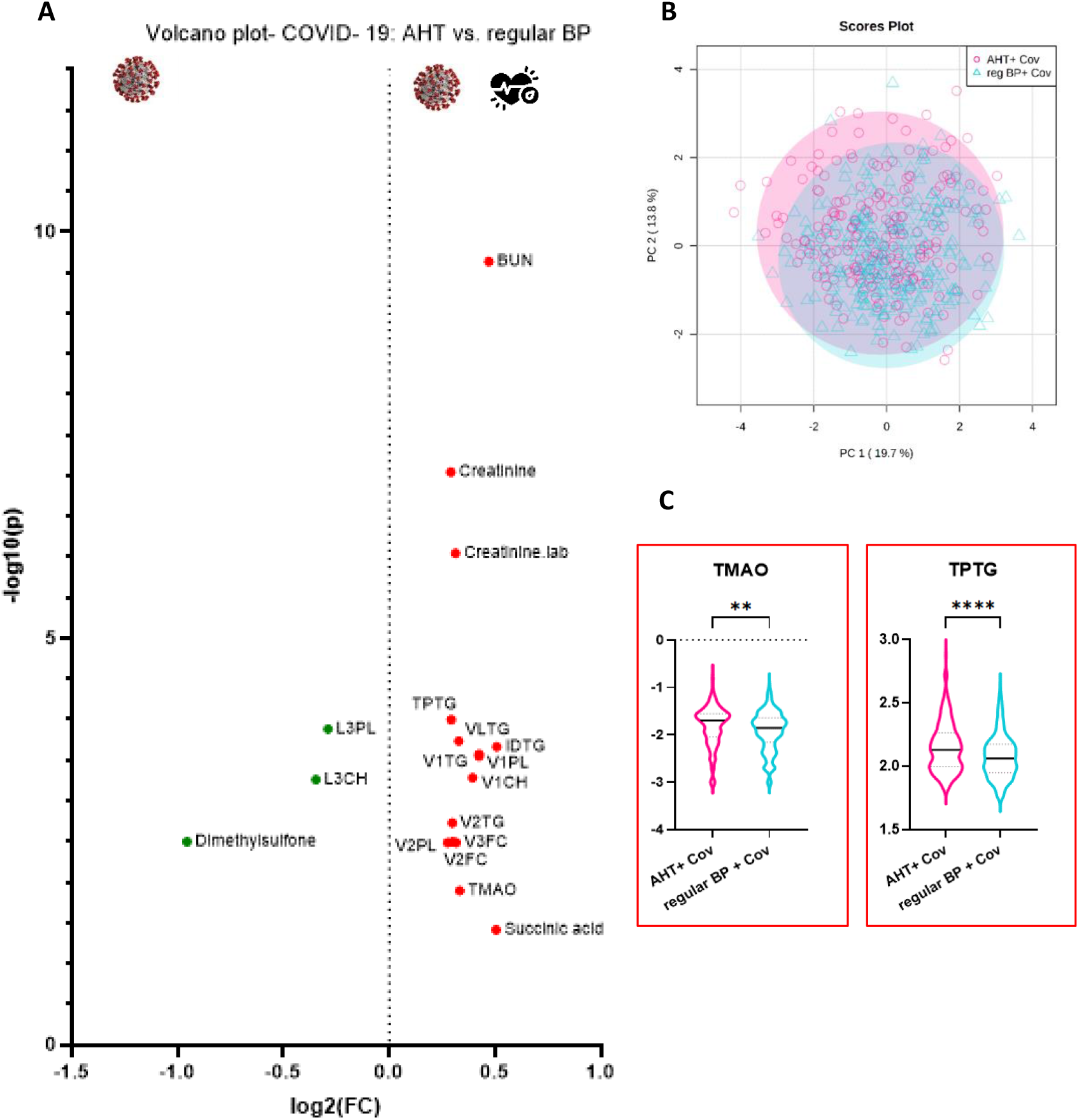
Metabolomic differences in normotensive and hypertensive COVID-19 patients. In (A) univariate analysis by means of volcano plot showing significant differences in normotensive and hypertensive COVID-19 patients (FC > 1.2, p < 0.05, FDR < 0.01): The red features are increased in the COVID-19 + AHT cohort, while the green ones are decreased in COVID-19 + AHT. (B) Multivariate analysis by means of principal component analysis (PCA) showing the distribution of both groups. (C) Violin plots showing significant alterations of the cardiovascular risk marker trimethyla-mine-N-oxide (TMAO) and TPTG. The y-axis shows the normalized concentration. AHT= Arterial hypertension, BP = blood pressure, BUN = Blood urea nitrogen, Creatinine.lab = Creatinine from laboratory report, Cov = COVID-19.

Similar metabolic alterations were observed in hospitalized COVID-19 + AHT patients (Fig. 6). This group also showed elevated renal values (BUN, creatinine) and a triglyceride-rich lipoprotein profile compared with normotensive hospitalized COVID-19 patients. However, the lipoprotein subclass distribution differed from the COVID-19 + AHT group in total in that HDL triglycerides were elevated in the hospitalized COVID-19 + AHT group. These subgroups were not conspicuous in the previous comparison (Fig. 5).

**Fig 6.**
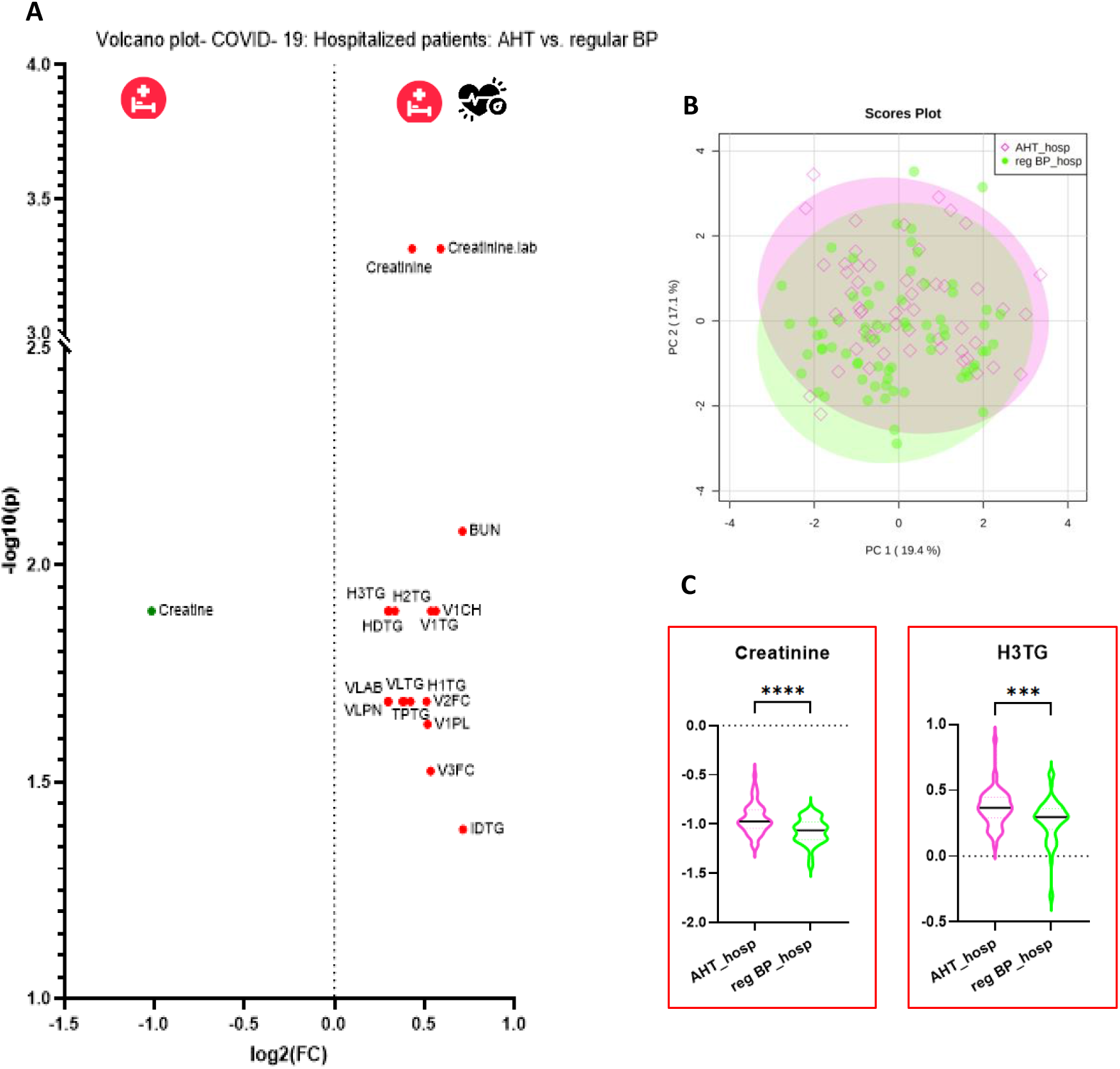
Metabolomic characteristics of hospitalized COVID-19 patients with AHT. In (A) univariate analysis by means of volcano plot showing significant differences between normotensive and hypertensive hospitalized COVID-19 patients (FC > 1.2, p < 0.05, FDR < 0.01): The red points show increased features in the hospitalized COVID-19 + AHT cohort, while the green one is decreased. (B) Multivariate analysis by means of principal component analysis (PCA) showing the distribution of both groups. In (C) violin plots show the most significant alterations of the NMR parameter Creatinine and H3TG. The y-axis shows the normalized concentration. AHT= Arterial hypertension, BP= blood pressure, hosp = hospitalized, BUN= Blood urea nitrogen, Creatinine.lab= Creatinine from laboratory report.

In particular, another volcano plot analysis was performed with 56 samples from 33 hospitalized COVID-19 + AHT patients and 71 samples from 38 normotensive hospitalized COVID-19 patients (Fig 6). Hereby, 17 features were elevated more than 1.2-fold, one metabolite (creatine) was decreased. Also, in this comparison, creatinine and BUN showed the highest significant alterations. The lipoprotein profile of the hospitalized COVID-19 + AHT cohort was characterized by increased total triglycerides and triglyceride subfractions (TPTG, VLTG, IDTG), increased VLDL particles in the total (VLPN), and in the less dense subfractions (VLDL-1, -2, -3). In addition, all HDL-triglyceride subfractions (HDTG, H1TG, H2TG, H3TG), except H4TG, were elevated in the hospitalized, hypertensive COVID-19 cohort.

Of the 134 hypertensive COVID-19 affected patients, 120 were treated with antihypertensives. 48 of these were treated with one medication, 72 with two or more. An overview of the medications of the hypertensive patients was given in Table 1. Antihypertensives indicated were angiotensin-converting-enzyme inhibitors (ACEI), angiotensin-1 receptor blockers (AT1RB), beta-blockers (BB), and calcium channel blockers (CCB). To determine any medication differences in metabolites and lipoproteins, the COVID-19 + AHT cohort was divided into subgroups according to their antihypertensive treatment. Patients treated with ACEI and/or AT1RB alone were grouped as the renin-angiotensin-aldosterone system inhibitor (RAASI) group, which included 75 samples. Hypertensive patients treated with BB and/or CCB formed the no RAASI group, which included 40 samples. The majority in this group were treated with BB (29 samples). 75 samples were from patients treated with different combinations of ACEI/AT1RB and BB and/or CCB. The 26 samples from patients who were not taking antihypertensives according to metadata were excluded from this analysis. The groups were compared with univariate operations. Here, the RAASI-group showed improved renal values compared with the no RAASI group, in terms of decreased creatinine levels and higher glomerular filtration rate (GFR). This was not observed when patients were treated with a combination of RAASI and BB or CCB. In general, no significant alterations could be seen in the comparison of a combination of RAASI with BB/CCB versus the no RAASI group. Interestingly, increased VLDL particles (VLPN), consisting of the denser subgroups VLDL-4, were observed in the RAASI group. All significant changes are shown in the volcano plot in Fig 7.

**Fig 7.**
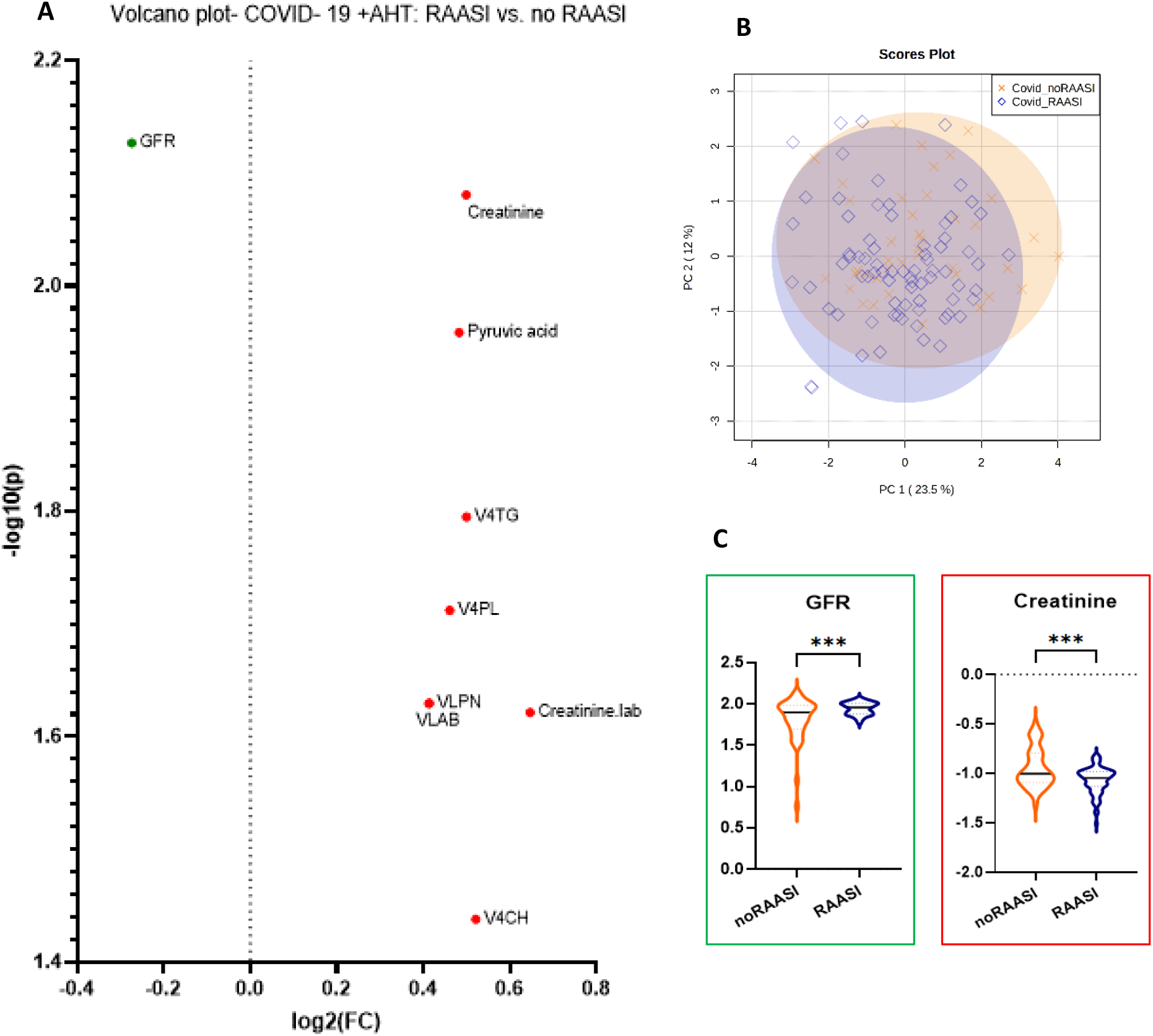
Hypertensive COVID-19 patients show improved kidney values under treatment with renin- angiotensin-aldoste-rone-inhibitors (RAASI) In (A) univariate comparison by means of volcano plot analysis (FC > 1.2, p < 0.05, FDR < 0.01): The red features correspond to increased values in the RAASI group, while the green ones correspond to decreased values. (B) Multivariate analysis by means of principal component analysis (PCA) showing the distribution of both groups. (C) Violin plots show significant alterations in the kidney parameters Creatinine and GFR = glomerular filtration rate. The y-axis shows the normalized concentration. Creatinine.lab= Creatinine from laboratory report.

### Hypertensive COVID-19 patients show higher inflammatory parameters

As a further univariate test method, an unpaired t-test was performed with the cohorts AHT versus COVID-19 + AHT. Another unpaired t-test was performed for the comparison COVID-19 with AHT versus regular BP. The p-value threshold was set at 0.05 (FDR < 0.01). These t-tests were performed for the already mentioned typical altered metabolites (glutamine, isoleucine, leucine, lysine, citric acid) and lipoproteins (HDL-4, TPCH, LDCH, SPC) in COVID-19 patients. The results are shown using violin plots in the supplementary data S20. It was additionally observed that hypertensive COVID-19 patients showed higher inflammation levels than normotensives. This was true for the NMR parameters Glyc, as well as the Glyc/SPC ratio, with the hypertensive COVID-19 group showing significantly higher values with p-values of p= 0.0014 for Glyc and p= 0.0058 for Glyc/SPC ratio. SPC was decreased in normotensive COVID-19 patients compared to the cohort without COVID-19 and therefore did not stand out as significant in this comparison, same for the laboratory value CRP, which was increased in normotensive and hypertensive COVID-19 patients. Violin plots for the mentioned inflammatory parameters are shown in Fig 8.

**Fig 8.**
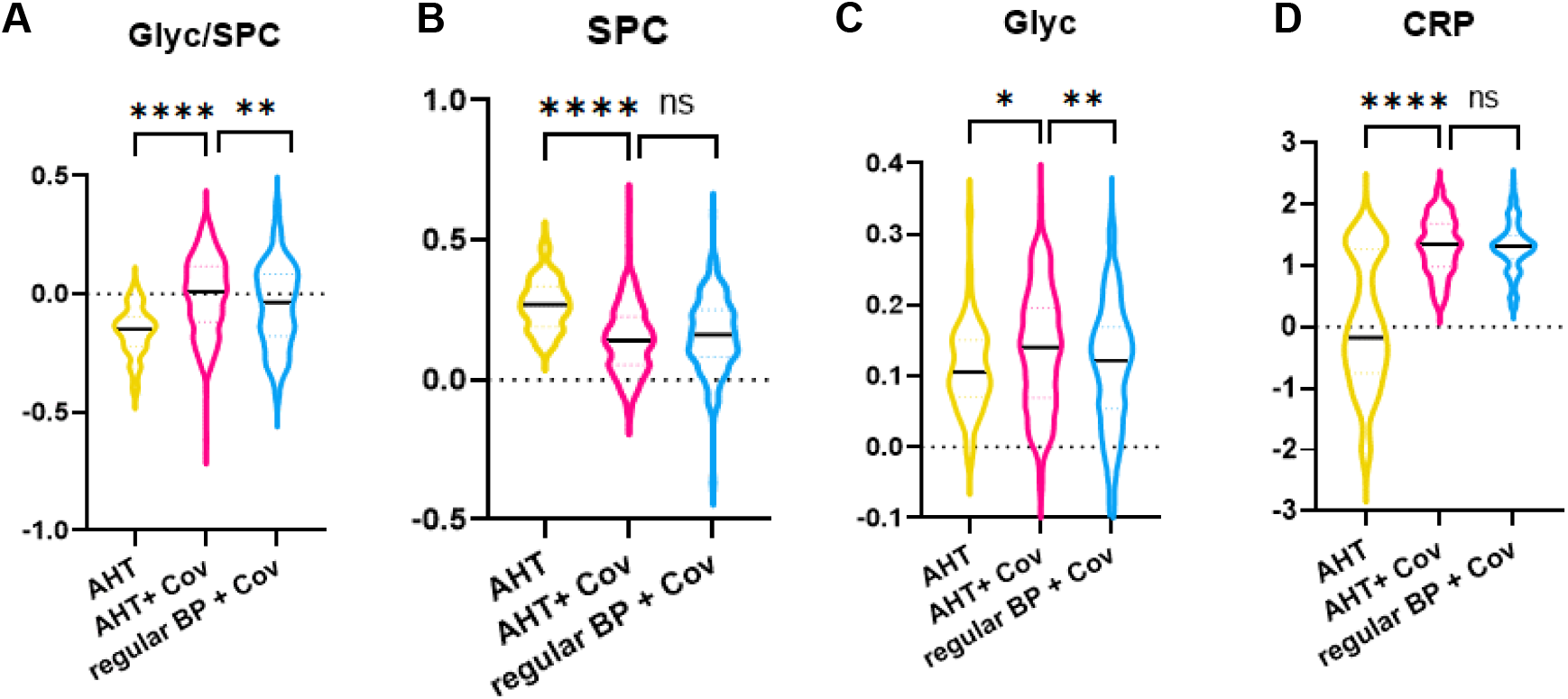
COVID-19 patients with AHT show higher inflammatory NMR parameters. Violin plots showing the inflammatory NMR parameters Glyc/SPC (A), SPC (B), Glyc (C), and routine lab CRP (D). Unpaired t-tests were performed for the comparison of AHT versus COVID-19 + AHT, and COVID-19 with and without AHT. Significant alterations are shown in form of asterisks: ns (not significant), p < 0.05 (*), p < 0.01 (**), p < 0.001 (***), p < 0.0001 (****). The y-axis shows the normalized concentration. AHT= Arterial hypertension, Cov= COVID-19, BP= Blood pressure, CRP= C-reactive protein.

## Discussion

In this study, we analyzed a COVID-19 cohort with 509 serum samples from 329 patients, which were treated mainly in an outpatient care setting. In our analyses, we focused on 134 patients (216 samples), which were additionally affected by arterial hypertension. To the best of our knowledge, this is the first study to investigate the effect of AHT on COVID-19 in the context of NMR-based metabolomics. We could show that the metabolic changes of COVID-19 disease behave differently when pre-existing AHT is considered. Stratification for AHT has also an impact on differences detected between genders. Furthermore, it became clear that hypertensive COVID-19 patients might at higher risk for complications, shown by certain changes in lipoproteins, metabolites, and inflammatory parameters.

The Coronataxi study took place in Germany, where, according to the Robert-Koch-Institute, AHT prevalence is about one-third in women and men, respectively. In subjects older than 65 years prevalence is even two third for both genders. In our COVID-19 cohort AHT was also very present with 40.7%. Previously it has been shown that individuals with pre-existing cardiovascular disease have an altered metabolic profile, particularly with respect to lipoproteins. These include, among others, increased VLDL particles and decreased HDL levels (31, 35). Since hypertensive patients may thus have elevated VLDL levels, they obviously drop out in the univariate statistics when COVID-19 patients with hypertension are compared with controls with hypertension, leading to the hypothesis that COVID-19, at least in milder courses, does not cause an increase in VLDL and its subgroups, as well as in the ABA1 ratio. However, what does suggest dyslipidemia in COVID-19-affected individuals despite this are the decreased levels in the HDL fractions. In particular, the decreased levels in the dense HDL-4 particles are considered typical of COVID-19 in our cohort. HDL appears to correlate inversely with cardiovascular risk (36). However, there are discrepancies in the extent to which the various HDL subgroups are involved in this process (37). Furthermore, since different methods are used to measure HDL subgroups, it is not always easy to compare the literature in this regard. Small HDL particles, such as HDL-4 (corresponding to HDL-3 in ultracentrifugation (38)), have been shown in some studies to be risk factors for cardiovascular disease (39). On the other hand, the protective and antioxidant effects on the endothelium (40) and the fibrinolytic properties of denser HDL particles had been described. (41). Apolipoproteins A1 and A2 each play a major role, as well as inverse correlating markers for cardiovascular risk (42). Adjacent to cardiovascular disease, HDL has a more clear relationship with the immune system (43, 44) and changes in its composition and function during an acute phase reaction (45). This is characterized by a decrease in apolipoprotein A1 and HDL cholesterol, which was evident in our COVID-19 cohort where we identified a decrease in total (TPA1 and TPCH, respectively) and HDL groups HDA1 and H4A1, and HDCH and H4CH, respectively. Thus, it can be inferred that the acute-phase response induced by COVID-19 may cause dysfunctionality of HDL particles, leading to loss of atheroprotective properties (46). In addition to the decrease in HDL-particles, several LDL-fractions also showed a decrease. This is consistent with other studies (8, 14). Furthermore, a correlation between increasing disease severity and a decrease in LDL cholesterol, as well as an inverse correlation between LDL cholesterol and CRP could be observed (47, 48). Possible reasons for this include impaired liver function, altered lipid metabolism under the influence of cytokines, and the breakdown of lipids by free radicals (47).

### General metabolic differences in COVID-19 patients

Regarding metabolites, COVID-19 characteristics include decreases in amino acids glutamine, isoleucine, leucine, and lysine. This indicates an altered energy metabolism in the course of hypoxemia (49). Decreased glutamine correlated inversely with CRP, lactate dehydrogenase (LDH), and partial pressure of oxygen in this study (49). In addition, glutamine fulfills important functions in the body, e.g. as a reactant for glutathione biosynthesis or as an energy supplier for immune cells (50). The reduction in the other amino acids mentioned could indicate increased amino acid catabolism, possibly as a result of temporary fasting during acute infection, as of our COVID-19 cohort, 22% reported loss of appetite and 9% nausea. Another metabolite that is decreased in COVID-19 is citric acid. This Krebs cycle metabolite also correlated inversely with CRP and LDH in one study (51).

### The role of AHT on previous findings

Comparing the AHT-control cohort with the COVID-19 + AHT cohort, the acute phase protein CRP showed an increase, which is not surprising, as well as the NMR-based inflammatory parameter Glyc/SPC. This ratio was also elevated in another study of COVID-19 patients (28). Glycoproteins (Glyc) represent the acute phase reaction (23). Supramolecular Phospholipid Composite signal (SPC) reproduces lipoproteins, especially apolipoproteins and HDL (28). Another prominent parameter in the COVID-19 cohort was phenylalanine. It has already been mentioned that this amino acid is elevated in COVID-19 patients and also correlates with a more severe progression (17). Even before the COVID-19 pandemic, phenylalanine was shown to be elevated in severe infections and associated with increased mortality, intensive care unit (ICU) treatment, and CRP levels (52). The elevated phenylalanine levels may be related to impaired liver or kidney function, as the hydroxylation of phenylalanine to tyrosine occurs in these organs (53). At the same time, tyrosine levels are reduced in the COVID-19 + AHT cohort, raising further suspicion that this process may be impaired, as it is also the case in patients with severe liver injury (54), and chronic kidney failure (55). The increased levels of glutamic acid in the COVID-19 patients may result from an increased supply from glutamine, for the synthesis of glutathione as an antioxidant or as an alternative energy resource for gluconeogenesis (56). Furthermore, COVID-19 patients exhibit elevated levels of the Krebs cycle intermediate succinic acid. This is possibly an expression of a hypoxic metabolic state, since succinic acid is able to stabilize hypoxia inducible factor-1α (HIF-1α) (57) and also to force the interleukin production (58).

### Gender differences in COVID-19

Male sex is a known risk factor for severe COVID-19 progression and complications (59, 60). In our gender comparison of COVID-19 individuals, several parameters could be identified as markers for an unfavorable disease course. These included elevated ketone bodies (acetone, acetoacetic acid, 3-hydroxybutyric acid) and marked dyslipidemia (16). The latter was indicated by increased VLDL particles in males and decreased HDL levels compared to female SARS-CoV-2 infected individuals. From this, one could deduce that the risk factor “male gender” can also be observed in the metabolome. However, a different picture emerged when the AHT factor was also included in this comparison. Only isolated VLDL particles then showed increased values. Ketone bodies, Glyc/SPC and HDL particles were also no longer found to be significantly altered. The sex differences were mainly related to an increase in various LDL fractions in women, as well as an increase in the BCAAs leucine and isoleucine, citric acid, and three VLDL particles. The elevated levels of dimethyl sulfone in our study could be attributed to two patients with very high levels and is possibly the result of a specific diet. However, considering sex and AHT, there were also some common features that, although not specific for COVID-19, emerged as typical metabolic profile in with SARS-CoV-2 infected individuals. These included increases in CRP and Glyc/SPC, and decreases in glutamine, isoleucine, leucine, lysine, citric acid, HDL-4 subsets, total cholesterol, LDL-cholesterol, and SPC. This is consistent with other metabolomics studies (8, 9, 13, 14) on COVID-19 but gives a finer picture of metabolic characteristics.

### AHT as risk factor in COVID-19: What can be seen in the metabolome?

AHT has also previously been cited as a risk factor for severe COVID-19 progression (4) and is also associated with an increased risk of various cardiovascular complications, such as heart failure or acute coronary syndrome (61). In addition to the already discussed changes in the lipoproteome in cardiac pre-diseased patients, the metabolite trimethylamine-N-oxides (TMAO) should also be mentioned. This metabolite, which is synthesized by intestinal bacteria, is considered a cardiovascular risk marker due to its proartherosclerotic effects. (62).

As expected, also in our cohort with mostly milder courses, hypertensive COVID-19 patients showed an increase in the mentioned cardiovascular risk markers compared with normotensive COVID-19 affected individuals. Interestingly, the ABA1 ratio did not stand out as significantly elevated in hypertensive patients in our cohort. Furthermore, signs of impaired renal function were clearly observed in hypertensive COVID-19 patients. However, these markers are less an expression of COVID-19 but rather due to AHT as such. Thus, when comparing the AHT-cohorts with and without COVID-19, there was no increase in these parameters and thus no further deterioration of renal function during acute infection. However, an indication of renal impairment in the COVID-19 + AHT cohort, is the observation that tyrosine was found to be significantly decreased in hypertensives only in the comparison AHT with and without COVID-19. The triglyceride-rich lipoprotein profile also did not appear to be linked to COVID-19. Succinic acid, which was elevated in hypertensives, could represent a more severe COVID-19 course as an expression of a hypoxic metabolic state (57). TMAO, which was increased in hypertensive COVID-19 patients, was also associated with a more severe COVID-19 course in patients with pre-existing diabetes mellitus or cardiovascular diseases in one study (63). Also, COVID-19 patients with pre-existing AHT showed higher inflammatory values in terms of Glyc and Glyc/SPC ratio in our study compared to patients without pre-existing AHT. Comparing hospitalized COVID-19 patients with and without AHT, it was also not surprising that COVID-19 patients with AHT had higher renal values (creatinine, BUN), as well as a lipoprotein profile rich in triglycerides.

### Influence of various antihypertensives on COVID-19 + AHT patients

With regard to antihypertensive treatment, the use of angiotensin converting enzyme inhibitors (ACEI) and angiotensin II receptor blockers (ARB) was discussed at the beginning of the pandemic. It was suspected that these classes of drugs, by increasing the expression of ACE-2 receptors, would lead to increased viral uptake into cells and thus to a worse course of COVID-19 disease (64). However, it has now been shown that treatment with ACEIs and ARBs is not associated with a more severe disease course or increased mortality, but rather the opposite, and blood pressure control is an important factor in COVID-19 treatment (65, 66). Also in our results, COVID-19 + AHT patients, treated with this group of drugs had lower mean levels of renal creatinine and a higher GFR than patients treated with other antihypertensives. However, with the use of RAASI, there was also an increase in some VLDL particles. This could be due to a higher number of multimorbid patients in this group.

## Conclusion

Here, we presented an NMR-based study examining serum from a large cohort of outpatients and hospitalized COVID-19 patients. Extensive meta-data allowed us to relate to common laboratory parameters, as well as to include in our analyses the important comorbidity AHT, which occurs with high prevalence. This provides a much finer picture of the metabolic fingerprint of COVID-19 disease. The extensive quantification of metabolites and lipoproteins with an uncomplicated and rapid measurement method such as NMR opens many possibilities in the field of pathophysiology research and risk stratification, which has not yet been conclusively elucidated in COVID-19.

We are aware that the statistical power is diminished by larger differences in cohort sizes and by the fact that no comparisons of laboratory parameters and parameter from the B.I. PACS™ report could be included in the comparison of the COVID-19 patient cohort with the healthy control group from Bruker (HC).

## Data Availability

All data produced in the present study are available upon reasonable request to the authors.

## Abbreviations

Name: Extended name
ACEI: Angiotensin-converting-enzyme inhibitor
AHT: Arterial hypertension
AT1RB: Angiotensin-1 receptor blockers
AUC: Area under the curve
BB: Beta blockers
BCAA: Branched chained amino acids (leucine, isoleucine, valine)
BP: Blood pressure
BUN: Blood urea nitrogen
CCB: Calcium channel blockers
Creatinine.lab: Creatinine (laboratory report)
CRP: C-reactive protein
FC: Fold change
FDR: False Discovery rate
GFR: Glomerular filtration rate
GGT: Gamma-glutamyl transferase
Glyc: Glycoproteins
GOT: Glutamate-oxalacetate-transferase
HC: Healthy cohort
Leuk: Leucozytes
LDH: Lactat dehydrogenase
NMR: Nuclear magnetic resonance
SPC: Supramolecular phospholipid composite
T2DM: Diabetes mellitus Type 2
TMAO: Trimethylamine-N-oxide
PACS: Post-acute-covid-syndrom
PCA: Principal component analysis
RAASI: Renin-angiotensin-aldosterone-system inhibitors

## Funding

This project was supported by the Deutsche Forschungsgemeinschaft (DFG, German Research Foundation)-project number 374031971-TRR 240.

## Acknowledgments

We thank the Werner Siemens Imaging Center with the Chair of Department Prof. Dr. Pichler for the opportunity to perform this research. In addition, we greatly thank Karin Tarbet from the Coronataxi-team for her great work and effort and the medical technicians Petra Klöters-Plachky and Alexandra Hof for their biobanking work. Further, we want to thank the members of CRC/TR240 project Z03 (Cornelia Fiessler, Anna Grau, Steffi Jiru-Hillmann, Kirsten Haas) from the institute for clinical epidemiology and biometry, University Hospital Würzburg, Würzburg, Germany and Frederic Emschermann from the Department of Internal Medicine III, Cardiology and Angiology, University Hospital Tübingen, Tübingen, Germany for the good cooperation. We also thank our technical assistants Miriam Owczorz and Daniele Bucci for the support during sample preparation and NMR measurements.

## Notes

### Competing Interest Statement

CT and GB report a research grant by Bruker BioSpin GmbH.

### Author Declarations

This study was conducted according to the Declaration of Helsinki and was approved by the Ethics Committees of Heidelberg Medical University (protocol code S/324/2020), University Hospital Tuebingen (protocol code 141/2018BO2) and University of Wuerzburg (protocol code 52/18).

## References

1. Zhou F, Yu T, Du R, Fan G, Liu Y, Liu Z, et al. Clinical course and risk factors for mortality of adult inpatients with COVID-19 in Wuhan, China: a retrospective cohort study. The Lancet. 2020;395(395):1054–62.

2. Scudiero F, Silverio A, Di Maio M, Russo V, Citro R, Personeni D, et al. Pulmonary embolism in COVID-19 patients: prevalence, predictors and clinical outcome. Thrombosis Research. 2021;198:34–9.

3. Farshidfar F, Koleini N, Ardehali H. Cardiovascular complications of COVID-19. JCI Insight. 2021;6(13).

4. Gao Y-d, Ding M, Dong X, Zhang J-j, Kursat Azkur A, Azkur D, et al. Risk factors for severe and critically ill COVID-19 patients: A review. Allergy. 2021;76(76):428–55.

5. Jin J-M, Bai P, He W, Wu F, Liu X-F, Han D-M, et al. Gender Differences in Patients With COVID-19: Focus on Severity and Mortality. Frontiers in Public Health. 2020;8.

6. Letertre MPM, Giraudeau P, de Tullio P. Nuclear Magnetic Resonance Spectroscopy in Clinical Metabolomics and Personalized Medicine: Current Challenges and Perspectives. Front Mol Biosci. 2021;8:698337-.

7. Pang H, Jia W, Hu Z. Emerging applications of metabolomics in clinical pharmacology. Clinical Pharmacology & Therapeutics. 2019;106(106):544–56.

8. Bruzzone C, Bizkarguenaga M, Gil-Redondo R, Diercks T, Arana E, García de Vicuña A, et al. SARS-CoV-2 Infection Dysregulates the Metabolomic and Lipidomic Profiles of Serum. iScience. 2020;23(23):101645.

9. Baranovicova E, Bobcakova A, Vysehradsky R, Dankova Z, Halasova E, Nosal V, et al. The Ability to Normalise Energy Metabolism in Advanced COVID-19 Disease Seems to Be One of the Key Factors Determining the Disease Progression—A Metabolomic NMR Study on Blood Plasma. Applied Sciences. 2021;11(11):4231.

10. Dierckx T, van Elslande J, Salmela H, Decru B, Wauters E, Gunst J, et al. The metabolic fingerprint of COVID-19 severity. medRxiv. 2020:2020.11.09.20228221.

11. Ghini V, Meoni G, Pelagatti L, Celli T, Veneziani F, Petrucci F, et al. Profiling metabolites and lipoproteins in COMETA, an Italian cohort of COVID-19 patients. PLOS Pathogens. 2022;18(18):e1010443.

12. Schmelter F, Foeh B, Mallagaray A, Rahmoeller J, Ehlers M, Lehrian S, et al. Metabolic markers distinguish COVID-19 from other intensive care patients and show potential to stratify for disease risk. medRxiv. 2021:2021.01.13.21249645.

13. Lodge S, Nitschke P, Kimhofer T, Coudert JD, Begum S, Bong S-H, et al. NMR Spectroscopic Windows on the Systemic Effects of SARS-CoV-2 Infection on Plasma Lipoproteins and Metabolites in Relation to Circulating Cytokines. Journal of Proteome Research. 2021;20(20):1382–96.

14. Kimhofer T, Lodge S, Whiley L, Gray N, Loo RL, Lawler NG, et al. Integrative Modeling of Quantitative Plasma Lipoprotein, Metabolic, and Amino Acid Data Reveals a Multiorgan Pathological Signature of SARS-CoV-2 Infection. Journal of proteome research. 2020;19(19):4442--54, pmid = 32806897.

15. Aboelela MI, Abd Elmajeed AD, Abd Elmaksoud H, Sayed AA. AStudy on APO B100/APO A1 Ratio as A Predictive Parameter for Assessmentof CAD Risk in Uncontrolled Type 2 Egyptian Diabetic Patients. The Egyptian Journal of Hospital Medicine. 2014;54(54):62–70.

16. Rendeiro AF, Vorkas CK, Krumsiek J, Singh H, Kapatia S, Cappelli LV, et al. Metabolic and immune markers for precise monitoring of COVID-19 severity and treatment. medRxiv. 2021:2021.09.05.21263141.

17. Luporini RL, Pott-Junior H, Di Medeiros Leal MCB, Castro A, Ferreira AG, Cominetti MR, et al. Phenylalanine and COVID-19: Tracking disease severity markers. International Immunopharmacology. 2021;101:108313.

18. Feingold KR. Lipid and Lipoprotein Levels in Patients with COVID-19 Infections. In: Feingold KR, Anawalt B, Boyce A, Chrousos G, de Herder WW, Dhatariya K, et al., editors. Endotext. South Dartmouth (MA): MDText.com, Inc. Copyright © 2000-2022, MDText.com, Inc.; 2022.

19. Meoni G, Ghini V, Maggi L, Vignoli A, Mazzoni A, Salvati L, et al. Metabolomic/lipidomic profiling of COVID-19 and individual response to tocilizumab. PLOS Pathogens. 2021;17(17):e1009243.

20. Nalbandian A, Sehgal K, Gupta A, Madhavan MV, McGroder C, Stevens JS, et al. Post-acute COVID-19 syndrome. Nature Medicine. 2021;27(27):601–15.

21. Holmes E, Wist J, Masuda R, Lodge S, Nitschke P, Kimhofer T, et al. Incomplete Systemic Recovery and Metabolic Phenoreversion in Post-Acute-Phase Nonhospitalized COVID-19 Patients: Implications for Assessment of Post-Acute COVID-19 Syndrome. Journal of proteome research. 2021;20(20):3315--29, pmid = 34009992.

22. Liptak P, Baranovicova E, Rosolanka R, Simekova K, Bobcakova A, Vysehradsky R, et al. Persistence of Metabolomic Changes in Patients during Post-COVID Phase: A Prospective, Observational Study. Metabolites. 2022;12(12):641.

23. Otvos JD, Shalaurova I, Wolak-Dinsmore J, Connelly MA, Mackey RH, Stein JH, et al. GlycA: A Composite Nuclear Magnetic Resonance Biomarker of Systemic Inflammation. Clinical Chemistry. 2015;61(61):714–23.

24. Fuertes-Martín R, Taverner D, Vallvé J-C, Paredes S, Masana L, Correig Blanchar X, et al. Characterization of 1H NMR Plasma Glycoproteins as a New Strategy To Identify Inflammatory Patterns in Rheumatoid Arthritis. Journal of Proteome Research. 2018;17(17):3730–9.

25. Nitschke P, Lodge S, Kimhofer T, Masuda R, Bong S-H, Hall D, et al. J-Edited DIffusional Proton Nuclear Magnetic Resonance Spectroscopic Measurement of Glycoprotein and Supramolecular Phospholipid Biomarkers of Inflammation in Human Serum. Analytical Chemistry. 2022;94(94):1333–41.

26. Otvos JD, Guyton JR, Connelly MA, Akapame S, Bittner V, Kopecky SL, et al. Relations of GlycA and lipoprotein particle subspecies with cardiovascular events and mortality: A post hoc analysis of the AIM-HIGH trial. Journal of Clinical Lipidology. 2018;12(12):348-55.e2.

27. Chiesa ST, Charakida M, Georgiopoulos G, Roberts JD, Stafford SJ, Park C, et al. Glycoprotein Acetyls: A Novel Inflammatory Biomarker of Early Cardiovascular Risk in the Young. Journal of the American Heart Association. 2022;11(11):e024380.

28. Lodge S, Nitschke P, Kimhofer T, Wist J, Bong S-H, Loo RL, et al. Diffusion and Relaxation Edited Proton NMR Spectroscopy of Plasma Reveals a High-Fidelity Supramolecular Biomarker Signature of SARS-CoV-2 Infection. Analytical Chemistry. 2021;93(93):3976–86.

29. Ameta K, Gupta A, Kumar S, Sethi R, Kumar D, Mahdi AA. Essential hypertension: A filtered serum based metabolomics study. Scientific Reports. 2017;7(7):2153.

30. Deng Y, Huang C, Su J, Pan C-W, Ke C. Identification of biomarkers for essential hypertension based on metabolomics. Nutrition, Metabolism and Cardiovascular Diseases. 2021;31(31):382–95.

31. Nayak P, Panda S, Thatoi PK, Rattan R, Mohapatra S, Mishra PK. Evaluation of Lipid Profile and Apolipoproteins in Essential Hypertensive Patients. J Clin Diagn Res. 2016;10(10):BC01–BC4.

32. Würtz P, Havulinna AS, Soininen P, Tynkkynen T, Prieto-Merino D, Tillin T, et al. Metabolite Profiling and Cardiovascular Event Risk. Circulation. 2015;131(131):774–85.

33. Del Coco L, Vergara D, De Matteis S, Mensà E, Sabbatinelli J, Prattichizzo F, et al. NMR-Based Metabolomic Approach Tracks Potential Serum Biomarkers of Disease Progression in Patients with Type 2 Diabetes Mellitus. Journal of Clinical Medicine. 2019;8(8):720.

34. Lim A, Hippchen T, Unger I, Heinze O, Welker A, Kräusslich H-G, et al. An Outpatient Management Strategy Using a Coronataxi Digital Early Warning System Reduces Coronavirus Disease 2019 Mortality. Open Forum Infectious Diseases. 2022;9(4).

35. Holmes MV, Millwood IY, Kartsonaki C, Hill MR, Bennett DA, Boxall R, et al. Lipids, Lipoproteins, and Metabolites and&#xa0;Risk of Myocardial Infarction and&#xa0;Stroke. Journal of the American College of Cardiology. 2018;71(71):620–32.

36. Rader DJ, Hovingh GK. HDL and cardiovascular disease. The Lancet. 2014;384(384):618–25.

37. Superko HR, Pendyala L, Williams PT, Momary KM, King SB, Garrett BC. High-density lipoprotein subclasses and their relationship to cardiovascular disease. Journal of Clinical Lipidology. 2012;6(6):496–523.

38. Gebhard C, Rhainds D, Tardif J-C. HDL and cardiovascular risk: is cholesterol in particle subclasses relevant? European Heart Journal. 2014;36(36):10–2.

39. Piko P, Kosa Z, Sandor J, Seres I, Paragh G, Adany R. The profile of HDL-C subfractions and their association with cardiovascular risk in the Hungarian general and Roma populations. Scientific Reports. 2022;12(12):10915.

40. De Souza JA, Vindis C, Nègre-Salvayre A, Rye K-A, Couturier M, Therond P, et al. Small, dense HDL 3 particles attenuate apoptosis in endothelial cells: pivotal role of apolipoprotein A-I. Journal of Cellular and Molecular Medicine. 2010;14(14):608–20.

41. Harbaum L, Ghataorhe P, Wharton J, Jiménez B, Howard LSG, Gibbs JSR, et al. Reduced plasma levels of small HDL particles transporting fibrinolytic proteins in pulmonary arterial hypertension. Thorax. 2019;74(74):380–9.

42. Florvall G, Basu S, Larsson A. Apolipoprotein A1 Is a Stronger Prognostic Marker Than Are HDL and LDL Cholesterol for Cardiovascular Disease and Mortality in Elderly Men. The Journals of Gerontology: Series A. 2006;61(61):1262–6.

43. Nazir S, Jankowski V, Bender G, Zewinger S, Rye K-A, van der Vorst EPC. Interaction between high-density lipoproteins and inflammation: Function matters more than concentration! Advanced Drug Delivery Reviews. 2020;159:94–119.

44. de la Llera Moya M, McGillicuddy FC, Hinkle CC, Byrne M, Joshi MR, Nguyen V, et al. Inflammation modulates human HDL composition and function in vivo. Atherosclerosis. 2012;222(222):390–4.

45. Bindu G H, Rao VS, Kakkar VV. Friend Turns Foe: Transformation of Anti-Inflammatory HDL to Proinflammatory HDL during Acute-Phase Response. Cholesterol. 2011;2011:274629.

46. Rosenson RS, Brewer HB, Ansell BJ, Barter P, Chapman MJ, Heinecke JW, et al. Dysfunctional HDL and atherosclerotic cardiovascular disease. Nature Reviews Cardiology. 2016;13(13):48–60.

47. Wei X, Zeng W, Su J, Wan H, Yu X, Cao X, et al. Hypolipidemia is associated with the severity of COVID-19. Journal of Clinical Lipidology. 2020;14(14):297–304.

48. Mahat RK, Rathore V, Singh N, Singh N, Singh SK, Shah RK, et al. Lipid profile as an indicator of COVID-19 severity: A systematic review and meta-analysis. Clinical Nutrition ESPEN. 2021;45:91–101.

49. Páez-Franco JC, Torres-Ruiz J, Sosa-Hernández VA, Cervantes-Díaz R, Romero-Ramírez S, Pérez-Fragoso A, et al. Metabolomics analysis reveals a modified amino acid metabolism that correlates with altered oxygen homeostasis in COVID-19 patients. Scientific Reports. 2021;11(11):6350.

50. de Oliveira DC, da Silva Lima F, Sartori T, Santos ACA, Rogero MM, Fock RA. Glutamine metabolism and its effects on immune response: molecular mechanism and gene expression. Nutrire. 2016;41(41):14.

51. Shi D, Yan R, Lv L, Jiang H, Lu Y, Sheng J, et al. The serum metabolome of COVID-19 patients is distinctive and predictive. Metabolism. 2021;118:154739.

52. Huang S-S, Lin J-Y, Chen W-S, Liu M-H, Cheng C-W, Cheng M-L, et al. Phenylalanine- and leucine-defined metabolic types identify high mortality risk in patients with severe infection. International Journal of Infectious Diseases. 2019;85:143–9.

53. Møller N, Meek S, Bigelow M, Andrews J, Nair KS. The kidney is an important site for <i>in vivo</i> phenylalanine-to-tyrosine conversion in adult humans: A metabolic role of the kidney. Proceedings of the National Academy of Sciences. 2000;97(97):1242–6.

54. Tessari P, Vettore M, Millioni R, Puricelli L, Orlando R. Effect of liver cirrhosis on phenylalanine and tyrosine metabolism. Current Opinion in Clinical Nutrition & Metabolic Care. 2010;13(13):81–6.

55. Boirie Y, Albright R, Bigelow M, Nair KS. Impairment of phenylalanine conversion to tyrosine inend-stage renal disease causing tyrosine deficiency. Kidney International. 2004;66(66):591–6.

56. Newsholme P, Procopio J, Lima MMR, Pithon-Curi TC, Curi R. Glutamine and glutamate— their central role in cell metabolism and function. Cell Biochemistry and Function. 2003;21(21):1–9.

57. Selak MA, Armour SM, MacKenzie ED, Boulahbel H, Watson DG, Mansfield KD, et al. Succinate links TCA cycle dysfunction to oncogenesis by inhibiting HIF-α prolyl hydroxylase. Cancer Cell. 2005;7(7):77–85.

58. Tannahill GM, Curtis AM, Adamik J, Palsson-McDermott EM, McGettrick AF, Goel G, et al. Succinate is an inflammatory signal that induces IL-1β through HIF-1α. Nature. 2013;496(496):238–42.

59. Biswas M, Rahaman S, Biswas TK, Haque Z, Ibrahim B. Association of Sex, Age, and Comorbidities with Mortality in COVID-19 Patients: A Systematic Review and Meta-Analysis. Intervirology. 2021;64(64):36–47.

60. Viveiros A, Rasmuson J, Vu J, Mulvagh SL, Yip CYY, Norris CM, et al. Sex differences in COVID-19: candidate pathways, genetics of ACE2, and sex hormones. American Journal of Physiology-Heart and Circulatory Physiology. 2021;320(320):H296–H304.

61. Lee CCE, Ali K, Connell D, Mordi IR, George J, Lang EM, et al. COVID-19-Associated Cardiovascular Complications. Diseases. 2021;9(9):47.

62. Papandreou C, Moré M, Bellamine A. Trimethylamine N-oxide in relation to cardiometabolic health—cause or effect? Nutrients. 2020;12(12):1330.

63. Terruzzi I, Senesi P. Does intestinal dysbiosis contribute to an aberrant inflammatory response to severe acute respiratory syndrome coronavirus 2 in frail patients? Nutrition. 2020;79-80:110996.

64. Khashkhusha TR, Chan JSK, Harky A. ACE inhibitors and COVID-19: We don’t know yet. J Card Surg. 2020;35(35):1172–3.

65. Sharma R, Kumar A, Majeed J, Thakur AK, Aggarwal G. Drugs acting on the renin– angiotensin–aldosterone system (RAAS) and deaths of COVID-19 patients: a systematic review and meta-analysis of observational studies. The Egyptian Heart Journal. 2022;74(74):64.

66. Lee T, Cau A, Cheng MP, Levin A, Lee TC, Vinh DC, et al. Angiotensin Receptor Blockers and Angiotensin-Converting Enzyme Inhibitors in COVID-19: Meta-analysis/Meta-regression Adjusted for Confounding Factors. CJC Open. 2021;3(3):965–75.

