## Supplementary data for "Stratification of hypertensive COVID-19 patients by quantitative NMR spectroscopy of serum metabolites, lipoproteins and inflammation markers"

Supplementary results:

Baseline characteristics (S1 Table):

Clinical metadata:

| **Characteristics** | **Total** | **Female** | **Male** |
|---|---|---|---|
| Gender, no. (%) | 329 | 174 (52,9) | 155 (47,1) |
| Age, median (IQR) | 54 (44- 64) | 52 (41- 63) | 58 (48- 65) |
| BMI, median (IQR) | 27.9 (24.4- 32.2) | 26.7 (22.6- 32) | 29 (25.9- 32.3) |

| **Disease severity** | **Total** | **Female** | **Male** |
|---|---|---|---|
| Hospital total, no. (%) | 71 (22.8) | 25 (35.2) | 46 (64.8) |
| Duration of stay, median (IQR) | 6 (3-8) | 4 (3-7) | 6 (4-8) |
| Oxygen demand, no. (%) | 48 (67.6) | 15 (31.2) | 33 (68.8) |
| ICU, no. (%) | 8 (11.3) | 3 (37.5) | 5 (62.5) |
| Death, no. (%) | 5 (1.5) | 3 (60.0) | 2 (40.0) |

Pre-existing diseases and medication:

| **Pre-existing disorders** | **Total (%)** | **Female (%)** | **Male (%)** |
|---|---|---|---|
| AHT | 134 (40.7) | 63 (47.0) | 71 (53.0) |
| CAD | 33 (10.0) | 9 (27.3) | 24 (72.7) |
| Chronic heart failure | 6 (1.8) | 4 (66.7) | 2 (33.3) |
| D.M. | 44 (13.4) | 20 (45.5) | 24 (54.5) |
| Asthma | 44 (13.4) | 31 (70.5) | 13 (29.5) |
| COPD | 12 (3.6) | 4 (33.3) | 8 (66.7) |
| OSAS | 22 (6.7) | 7 (31.8) | 15 (68.2) |
| Depression | 23 (7.0) | 15 (65.2) | 8 (34.8) |
| Cancer | 15 (4.6) | 7 (46.7) | 8 (53.3) |
| Cancer in past | 23 (7.0) | 14 (60.9) | 9 (39.1) |
| Stroke in the past | 16 (4.9) | 8 (50.0) | 8 (50.0) |
| Rheumatoid Arthritis | 10 (3.0) | 9 (90.0) | 1 (10.0) |
| Hypothyroidism | 46 (14.0) | 35 (76.1) | 11 (23.9) |

| | | | |
|---|---|---|---|
| Inflammatory bowel disease | 6 (1.8) | 5 (83.3) | 1 (16.7) |
| Thrombosis in the past | 27 (8.2) | 16 (59.3) | 11 (40.7) |
| Organ transplantation in the past | 3 (0.9) | 0 (0) | 3 (100) |
| Vaccination | 6 (1.8) | 4 (66.7) | 2 (33.3) |
| Pregnancy | 4 (1.2) | 4 (100) | 0 (0) |
| AF | 12 (3.6) | 6 (50.0) | 6 (50.0) |

| **Medication** | **Total (%)** | **Female (%)** | **Male (%)** |
|---|---|---|---|
| Beta-blockers | 61 (18.5) | 31 (50.8) | 30 (49.2) |
| Calcium channel blockers | 32 (9.7) | 14 (43.8) | 18 (56.3) |
| ACE- Inhibitors | 43 (13.1) | 17 (39.5) | 26 (60.5) |
| AT1-R-blockers | 62 (18.8) | 34 (54.8) | 28 (45.2) |
| L-thyroxin | 41 (12.5) | 31 (75.6) | 10 (24.4) |
| ASS | 46 (14.0) | 18 (31.9) | 28 (60.9) |
| Clopidogrel | 5 (1.5) | 0 (0) | 5 (100) |
| Citalopram/Sertraline | 13 (4.0) | 7 (53.8) | 6 (46.2) |
| Mirtazapine | 3 (0.9) | 2 (66.7) | 1 (33.3) |
| Venlafaxine | 2 (0.6) | 2 (100) | 0 (0) |
| Opipramol/ Amitriptyline | 5 (1.5) | 4 (80.0) | 1 (20.0) |
| Pantoprazole | 40 (12.2) | 21 (52.5) | 19 (47.5) |
| Statine | 46 (14.0) | 13 (28.3) | 33 (71.7) |
| Metformin | 20 (6.1) | 7 (35.0) | 13 (65.0) |
| Allopurinol | 14 (4.3) | 1 (7.1) | 13 (92.9) |
| Insulin | 9 (2.7) | 3 (33.3) | 6 (66.7) |
| Oral antidiabetics | 10 (3.0) | 1 (10.0) | 9 (90.0) |
| NOAC | 17 (5.2) | 5 (29.4) | 12 (70.6) |
| Cortisone | 7 (2.1) | 5 (71.4) | 2 (28.6) |
| Adalimumab | 2 (0.6) | 0 (0) | 2 (100) |
| Azathioprine | 1 (0.3) | 1 (100) | 0 (0) |
| Tacrolimus | 3 (0.9) | 3 (100) | 0 (0) |
| Rituximab | 1 (0.3) | 0 (0) | 1 (100) |
| Methotrexate | 5 (1.5) | 4 (80.0) | 1 (20.0) |
| Spray | 29 (8.8) | 16 (55.2) | 13 (44.8) |

Analysis COVID-19 vs. Healthy Control

Univariate Analysis Table (sorted by p-value) (S2 Table)

| Parameter | FC | log2(FC) | p.ajusted | -log10(p) |
|---|---|---|---|---|
| Glutamine | 0.6841 | -0.54772 | 1.03E-111 | 110.99 |
| TPA2 | 0.72538 | -0.46319 | 2.41E-94 | 93.619 |
| HDA2 | 0.75719 | -0.40128 | 1.01E-87 | 86.997 |
| TPA1 | 0.75174 | -0.4117 | 1.25E-83 | 82.902 |
| Citric acid | 0.62126 | -0.68673 | 9.49E-79 | 78.023 |
| Lysine | 0.65143 | -0.61831 | 6.31E-76 | 75.2 |
| HDA1 | 0.76073 | -0.39455 | 2.87E-73 | 72.542 |
| H3CH | 0.69877 | -0.51711 | 1.25E-70 | 69.903 |
| HDPL | 0.73918 | -0.43599 | 2.41E-66 | 65.618 |
| HDCH | 0.71989 | -0.47415 | 1.06E-64 | 63.976 |
| H4PL | 0.73369 | -0.44676 | 3.85E-64 | 63.414 |
| H3PL | 0.73583 | -0.44255 | 1.50E-61 | 60.823 |
| H4A1 | 0.76592 | -0.38474 | 1.28E-57 | 56.893 |
| H4A2 | 0.72183 | -0.47026 | 1.43E-51 | 50.846 |
| TPCH | 0.76278 | -0.39065 | 6.51E-51 | 50.187 |
| H4CH | 0.70392 | -0.50651 | 6.67E-51 | 50.176 |
| H3A1 | 0.77994 | -0.35857 | 6.17E-49 | 48.21 |
| H2A1 | 0.74449 | -0.42567 | 1.81E-48 | 47.743 |
| H3A2 | 0.74249 | -0.42955 | 1.22E-45 | 44.912 |
| H3FC | 0.65433 | -0.61191 | 1.86E-37 | 36.73 |
| H2CH | 0.70586 | -0.50255 | 9.65E-37 | 36.016 |
| LDPL | 0.76539 | -0.38574 | 2.91E-36 | 35.535 |
| H2A2 | 0.70813 | -0.49791 | 2.81E-34 | 33.551 |
| LDCH | 0.72038 | -0.47318 | 1.54E-33 | 32.812 |
| V5FC | 1.5719 | 0.65249 | 2.62E-33 | 32.581 |
| V1TG | 1.716 | 0.77907 | 2.52E-32 | 31.598 |
| V2FC | 1.5272 | 0.6109 | 5.97E-31 | 30.224 |
| H2FC | 0.67684 | -0.56312 | 1.14E-29 | 28.943 |
| V1CH | 1.5871 | 0.6664 | 2.43E-28 | 27.614 |
| VLPN | 1.3416 | 0.42398 | 8.23E-28 | 27.085 |
| VLAB | 1.3416 | 0.42397 | 8.23E-28 | 27.085 |
| V5TG | 1.2223 | 0.28965 | 2.34E-27 | 26.63 |
| L1FC | 0.75605 | -0.40345 | 2.52E-27 | 26.598 |
| H1FC | 0.66575 | -0.58695 | 3.20E-27 | 26.494 |
| H4FC | 0.7114 | -0.49127 | 1.22E-26 | 25.913 |
| H2PL | 0.75959 | -0.39671 | 2.32E-26 | 25.635 |
| L1CH | 0.75049 | -0.41409 | 2.75E-26 | 25.56 |
| L4PL | 0.65059 | -0.62017 | 2.19E-25 | 24.659 |
| V2TG | 1.4587 | 0.54466 | 4.45E-25 | 24.352 |
| Histidine | 0.74462 | -0.42542 | 7.27E-25 | 24.138 |
| Leucine | 0.80476 | -0.31336 | 1.78E-24 | 23.749 |
| TPTG | 1.368 | 0.45202 | 7.09E-24 | 23.15 |
| VLTG | 1.4276 | 0.51356 | 9.27E-24 | 23.033 |
| LDFC | 0.80783 | -0.30787 | 3.29E-23 | 22.483 |

| | | | | |
|---|---|---|---|---|
| Dimethylsulfone | 1.3473 | 0.43009 | 2.53E-22 | 21.597 |
| L4CH | 0.64172 | -0.63999 | 7.55E-22 | 21.122 |
| VLFC | 1.2819 | 0.3583 | 2.43E-20 | 19.614 |
| L4FC | 0.70237 | -0.50969 | 2.52E-20 | 19.598 |
| L1TG | 1.3178 | 0.39817 | 1.34E-18 | 17.873 |
| H1CH | 0.70565 | -0.50298 | 1.64E-18 | 17.784 |
| L4AB | 0.68527 | -0.54526 | 1.73E-18 | 17.763 |
| L3CH | 0.66506 | -0.58845 | 3.34E-18 | 17.476 |
| HDFC | 0.82447 | -0.27846 | 3.92E-18 | 17.407 |
| V3TG | 1.3283 | 0.40955 | 3.93E-18 | 17.405 |
| L1PL | 0.81302 | -0.29864 | 7.64E-18 | 17.117 |
| L5CH | 0.70177 | -0.51092 | 9.27E-18 | 17.033 |
| L3PL | 0.71021 | -0.49368 | 1.71E-17 | 16.767 |
| V1PL | 1.4874 | 0.57278 | 3.09E-17 | 16.51 |
| L4PN | 0.69663 | -0.52153 | 3.11E-17 | 16.507 |
| H1A2 | 0.66862 | -0.58074 | 8.87E-17 | 16.052 |
| Isoleucine | 0.82895 | -0.27064 | 1.07E-16 | 15.969 |
| VLPL | 1.2479 | 0.31956 | 1.38E-16 | 15.859 |
| H1PL | 0.72123 | -0.47146 | 2.22E-16 | 15.653 |
| V2CH | 1.2642 | 0.33818 | 5.44E-16 | 15.265 |
| ABA1 | 1.2089 | 0.27374 | 5.46E-16 | 15.263 |
| V3FC | 1.292 | 0.36957 | 1.65E-15 | 14.782 |
| L5FC | 0.75664 | -0.40233 | 3.26E-15 | 14.486 |
| L5PL | 0.72952 | -0.45499 | 2.15E-14 | 13.667 |
| V2PL | 1.2718 | 0.34689 | 3.49E-14 | 13.457 |
| VLCH | 1.2022 | 0.26572 | 7.09E-13 | 12.149 |
| L5PN | 0.76034 | -0.39529 | 7.64E-13 | 12.117 |
| Phenylalanine | 1.2936 | 0.37136 | 1.96E-12 | 11.707 |
| L6FC | 0.81138 | -0.30155 | 1.96E-12 | 11.707 |
| IDPL | 0.7708 | -0.37557 | 2.86E-12 | 11.543 |
| L5AB | 0.76712 | -0.38248 | 2.93E-12 | 11.533 |
| Trimethylamine-N-oxide | 0.64414 | -0.63456 | 1.23E-11 | 10.911 |
| L3FC | 0.78187 | -0.355 | 1.23E-11 | 10.911 |
| IDTG | 1.507 | 0.59166 | 5.26E-11 | 10.279 |
| L3AB | 0.78125 | -0.35614 | 2.16E-10 | 9.6655 |
| Acetone | 1.7595 | 0.8152 | 2.20E-10 | 9.6586 |
| L3PN | 0.78201 | -0.35474 | 2.47E-10 | 9.6067 |
| H1A1 | 0.73887 | -0.4366 | 2.80E-10 | 9.5529 |
| Ornithine | 0.79665 | -0.32799 | 3.01E-10 | 9.5218 |
| IDFC | 0.81631 | -0.29282 | 2.32E-05 | 4.634 |
| V1FC | 1.3128 | 0.3926 | 2.76E-05 | 4.5593 |
| N-Dimethylglycine | 2.3684 | 1.2439 | 6.50E-05 | 4.1873 |
| Glutamic acid | 1.2456 | 0.31688 | 0.00022808 | 3.6419 |
| 3-Hydroxybutyric acid | 1.3828 | 0.46758 | 0.00078785 | 3.1036 |
| Succinic acid | 1.2426 | 0.31342 | 0.03106 | 1.5078 |

Multivariate Analysis: COVID-19 vs. Healthy Control (S3)

PCA

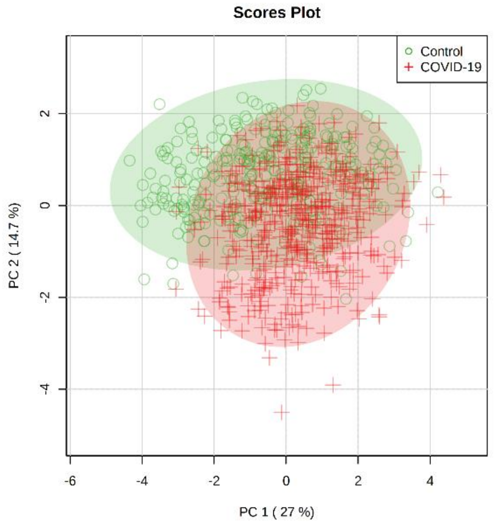


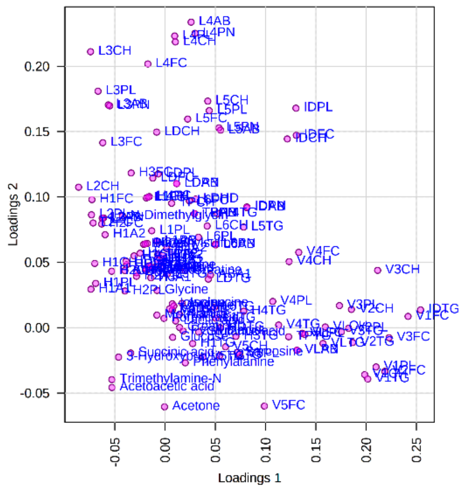


OPLS-DA

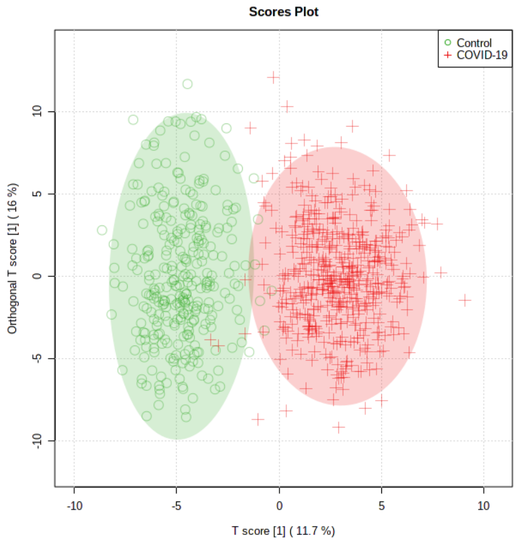


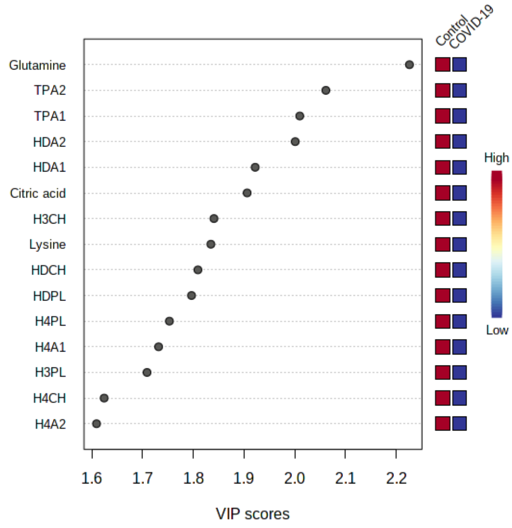


Analysis AHT vs. AHT + COVID-19

Univariate Analysis (sorted by p-value) (S4 Table)

| Parameter | FC | log2(FC) | p.ajusted | -log10(p) |
|---|---|---|---|---|
| Pyruvic acid | 0.34378 | -1.5404 | 3.42E-37 | 36.466 |
| CRP | 4.4144 | 2.1422 | 3.46E-32 | 31.461 |
| Lysine | 0.63898 | -0.64615 | 1.57E-22 | 21.803 |
| Glutamine | 0.75166 | -0.41185 | 3.22E-17 | 16.492 |
| H4A1 | 0.76258 | -0.39103 | 9.19E-14 | 13.037 |
| H4PL | 0.75366 | -0.40802 | 1.20E-13 | 12.922 |
| Alanine | 0.7504 | -0.41426 | 1.27E-13 | 12.895 |
| TPA1 | 0.81603 | -0.29331 | 2.47E-12 | 11.607 |
| Formic acid | 0.47993 | -1.0591 | 3.02E-12 | 11.519 |
| SPC | 0.76124 | -0.39358 | 1.33E-11 | 10.876 |

| | | | | |
|---|---|---|---|---|
| H4A2 | 0.73651 | -0.44123 | 3.52E-11 | 10.454 |
| Citric acid | 0.69175 | -0.53167 | 4.14E-11 | 10.383 |
| HDA1 | 0.82541 | -0.27681 | 8.26E-11 | 10.083 |
| Isoleucine | 0.66558 | -0.58731 | 2.03E-10 | 9.6925 |
| Leuk | 0.6963 | -0.52223 | 2.03E-10 | 9.6925 |
| TPA2 | 0.82631 | -0.27525 | 2.38E-10 | 9.6233 |
| Glyc/SPC | 1.4792 | 0.56478 | 4.70E-10 | 9.3278 |
| TPCH | 0.7705 | -0.37614 | 1.03E-09 | 8.9851 |
| H4CH | 0.71471 | -0.48458 | 1.36E-09 | 8.8666 |
| HDCH | 0.80149 | -0.31925 | 1.36E-09 | 8.8666 |
| L6CH | 0.7126 | -0.48884 | 3.25E-06 | 5.4875 |
| IDPL | 0.57773 | -0.79154 | 6.32E-06 | 5.199 |
| L6PL | 0.72773 | -0.45852 | 1.09E-05 | 4.962 |
| L5CH | 0.63683 | -0.65103 | 1.23E-05 | 4.909 |
| Leucine | 0.79357 | -0.33357 | 1.23E-05 | 4.909 |
| L6PN | 0.72834 | -0.45732 | 1.78E-05 | 4.7492 |
| L6AB | 0.73137 | -0.45133 | 2.22E-05 | 4.6532 |
| L6FC | 0.69577 | -0.52332 | 2.44E-05 | 4.6127 |
| Glutamic acid | 1.5094 | 0.59398 | 3.65E-05 | 4.4382 |
| Methionine | 0.76587 | -0.38482 | 5.96E-05 | 4.225 |
| L5PL | 0.67758 | -0.56154 | 7.55E-05 | 4.122 |
| Tyrosine | 0.78906 | -0.34179 | 0.00019964 | 3.6998 |
| Sarcosine | 0.60361 | -0.7283 | 0.00029499 | 3.5302 |
| L5FC | 0.7072 | -0.49982 | 0.00034 | 3.4685 |
| H1CH | 0.78061 | -0.35733 | 0.00043452 | 3.362 |
| H4FC | 0.7629 | -0.39043 | 0.00053694 | 3.2701 |
| IDTG | 0.58286 | -0.77878 | 0.00054617 | 3.2627 |
| LDCH | 0.75058 | -0.41393 | 0.00062519 | 3.204 |
| Histidine | 0.80503 | -0.31289 | 0.00069957 | 3.1552 |
| LDPL | 0.80256 | -0.31732 | 0.00082554 | 3.0833 |
| V1FC | 0.61237 | -0.70751 | 0.0010861 | 2.9641 |
| H3FC | 0.76657 | -0.38352 | 0.0010861 | 2.9641 |
| L4CH | 0.64315 | -0.63677 | 0.0013149 | 2.8811 |
| V1TG | 0.68941 | -0.53657 | 0.0018967 | 2.722 |
| L4PN | 0.6905 | -0.53429 | 0.0025942 | 2.586 |
| L4FC | 0.71626 | -0.48144 | 0.0025942 | 2.586 |
| L4AB | 0.68751 | -0.54056 | 0.0027858 | 2.5551 |
| V1PL | 0.67093 | -0.57576 | 0.0040313 | 2.3946 |
| LDPN | 0.82261 | -0.28172 | 0.004502 | 2.3466 |
| LDAB | 0.82262 | -0.28171 | 0.004502 | 2.3466 |
| L5PN | 0.71071 | -0.49266 | 0.0047199 | 2.3261 |
| L5AB | 0.70111 | -0.51229 | 0.00477 | 2.3215 |
| H1FC | 0.7784 | -0.36142 | 0.0049486 | 2.3055 |
| LDFC | 0.82459 | -0.27825 | 0.0053296 | 2.2733 |
| VLTG | 0.74069 | -0.43306 | 0.0055825 | 2.2532 |
| L1CH | 0.83058 | -0.26782 | 0.0064549 | 2.1901 |
| TPTG | 0.77155 | -0.37418 | 0.0072214 | 2.1414 |

| | | | | |
|---|---|---|---|---|
| VLPL | 0.79066 | -0.33887 | 0.0072214 | 2.1414 |
| L4PL | 0.67186 | -0.57377 | 0.0078989 | 2.1024 |
| L6TG | 0.82386 | -0.27952 | 0.02224 | 1.6529 |
| V1CH | 0.76374 | -0.38884 | 0.023154 | 1.6354 |
| IDFC | 0.75299 | -0.4093 | 0.023558 | 1.6279 |
| VLFC | 0.83132 | -0.26653 | 0.023837 | 1.6227 |
| Succinic acid | 1.3282 | 0.4095 | 0.02705 | 1.5678 |
| Phenylalanine | 1.2187 | 0.28535 | 0.04194 | 1.3774 |

Multivariate Analysis: AHT vs. COVID-19 + AHT (S5)

PCA

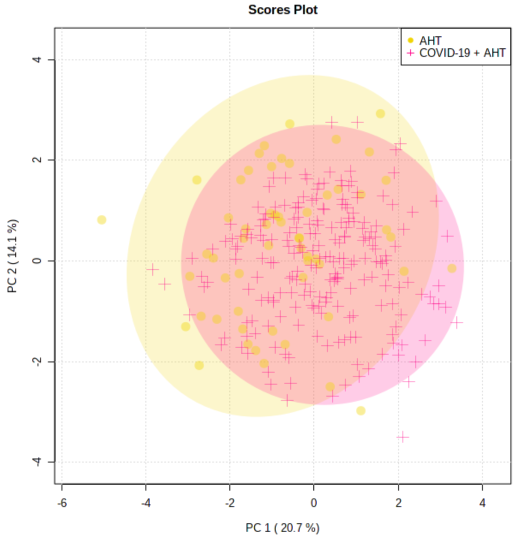


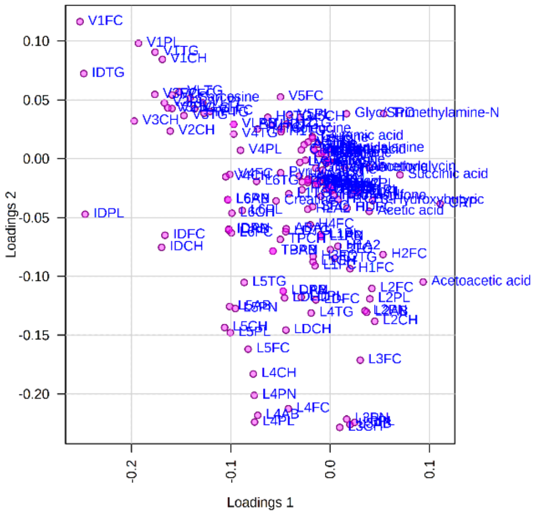


OPLS-DA

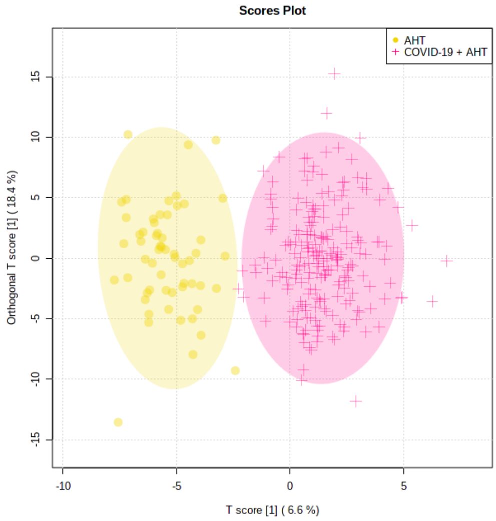


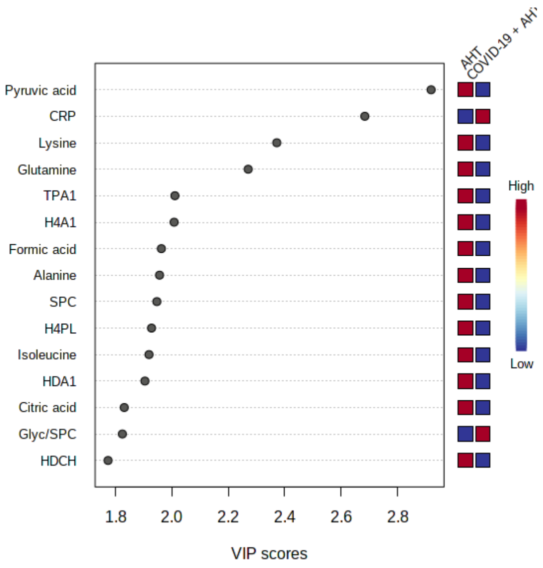

### Gender Analysis: COVID-19: Female vs. Male

Univariate Analysis (sorted by p-value) (S6 Table)

| Parameter | FC | log2(FC) | p.ajusted | -log10(p) |
|---|---|---|---|---|
| Kreatinin | 1.4708 | 0.55657 | 1.37E-41 | 40.865 |
| Creatinine | 1.4358 | 0.52186 | 1.28E-32 | 31.894 |
| Ferritin | 3.1099 | 1.6369 | 3.05E-32 | 31.516 |
| GGT | 2.162 | 1.1124 | 2.58E-21 | 20.589 |
| Harnstoff | 1.452 | 0.53803 | 1.05E-19 | 18.977 |
| Leucine | 1.297 | 0.37517 | 1.17E-17 | 16.932 |
| Valine | 1.2048 | 0.26876 | 2.11E-16 | 15.676 |
| H1PL | 0.72665 | -0.46066 | 1.25E-14 | 13.904 |
| H1CH | 0.74276 | -0.42903 | 2.25E-12 | 11.648 |
| Glyc/SPC | 1.2589 | 0.33212 | 6.51E-11 | 10.186 |
| H1FC | 0.73985 | -0.4347 | 6.69E-11 | 10.174 |
| H2CH | 0.82622 | -0.2754 | 9.13E-11 | 10.04 |
| Isoleucine | 1.2821 | 0.35848 | 1.07E-10 | 9.9699 |
| H1A1 | 0.72677 | -0.46042 | 1.10E-10 | 9.9567 |
| L1CH | 0.83057 | -0.26783 | 9.51E-10 | 9.0219 |
| H1A2 | 0.69659 | -0.52161 | 2.37E-09 | 8.626 |
| H3FC | 0.8012 | -0.31977 | 3.44E-08 | 7.4636 |
| L3FC | 0.8054 | -0.31222 | 3.24E-07 | 6.4889 |
| L3PL | 0.79081 | -0.33859 | 8.15E-07 | 6.0886 |
| H2FC | 0.82935 | -0.26994 | 1.36E-06 | 5.8654 |
| L2PL | 0.83079 | -0.26744 | 2.17E-06 | 5.6642 |
| L3CH | 0.77188 | -0.37355 | 2.71E-06 | 5.5678 |
| GOT | 1.2546 | 0.32724 | 3.83E-06 | 5.4167 |
| Creatine | 0.67374 | -0.56973 | 3.84E-06 | 5.4162 |
| L3AB | 0.81767 | -0.29041 | 8.30E-06 | 5.081 |
| L3PN | 0.81888 | -0.28827 | 9.59E-06 | 5.0181 |
| V2FC | 1.2606 | 0.33411 | 1.20E-05 | 4.9222 |
| V1PL | 1.2647 | 0.33877 | 1.56E-05 | 4.8081 |
| V1TG | 1.2463 | 0.31765 | 1.70E-05 | 4.7708 |
| VLTG | 1.2057 | 0.26991 | 2.66E-05 | 4.5754 |
| V3PL | 1.2113 | 0.27653 | 5.03E-05 | 4.2984 |
| V2PL | 1.2427 | 0.31349 | 6.74E-05 | 4.171 |
| L2CH | 0.82693 | -0.27415 | 0.00010996 | 3.9588 |
| V2TG | 1.237 | 0.30689 | 0.00022763 | 3.6428 |
| V1CH | 1.2212 | 0.28826 | 0.00022763 | 3.6428 |
| V3FC | 1.2045 | 0.26839 | 0.00024333 | 3.6138 |
| N-Dimethylglycine | 0.63067 | -0.66504 | 0.00026735 | 3.5729 |
| V1FC | 1.2921 | 0.36972 | 0.00065077 | 3.1866 |
| H1TG | 0.80807 | -0.30745 | 0.00075297 | 3.1232 |
| V2CH | 1.2189 | 0.28563 | 0.0011719 | 2.9311 |
| Acetoacetic acid | 1.8442 | 0.88301 | 0.0035446 | 2.4504 |
| IDTG | 1.269 | 0.34366 | 0.0040801 | 2.3893 |
| Acetone | 1.4223 | 0.50819 | 0.0046386 | 2.3336 |

| | | | | |
|---|---|---|---|---|
| 3-Hydroxybutyric acid | 1.6625 | 0.73332 | 0.0059167 | 2.2279 |
| Succinic acid | 1.4952 | 0.58032 | 0.0071047 | 2.1485 |

### Multivariate Analysis: COVID-19: Male vs. Female (S7)

#### PCA

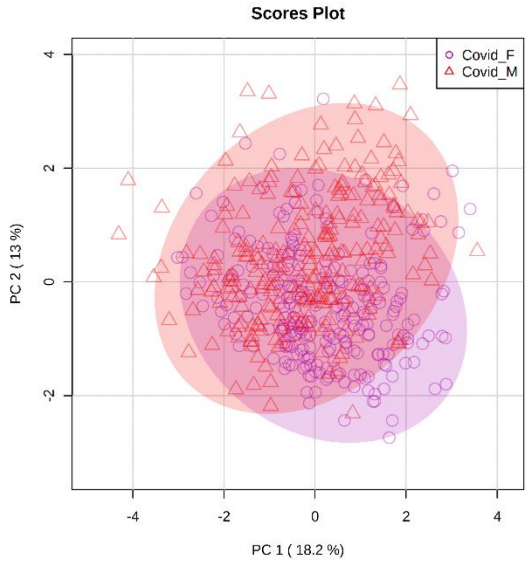


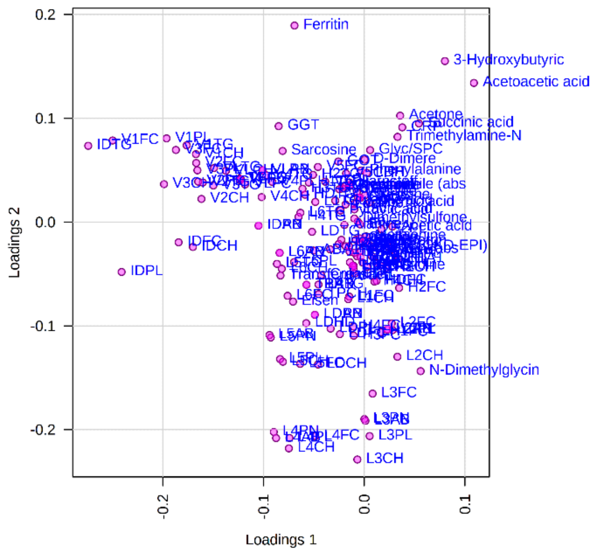


#### OPLS-DA

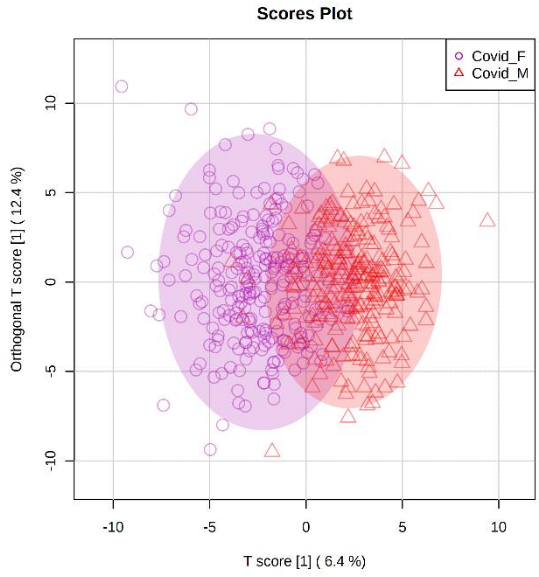


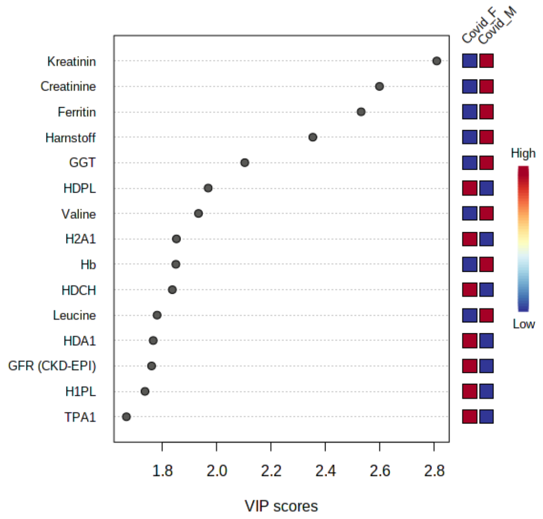

OPLS-DA (Creatinine, Kreatinin excluded)

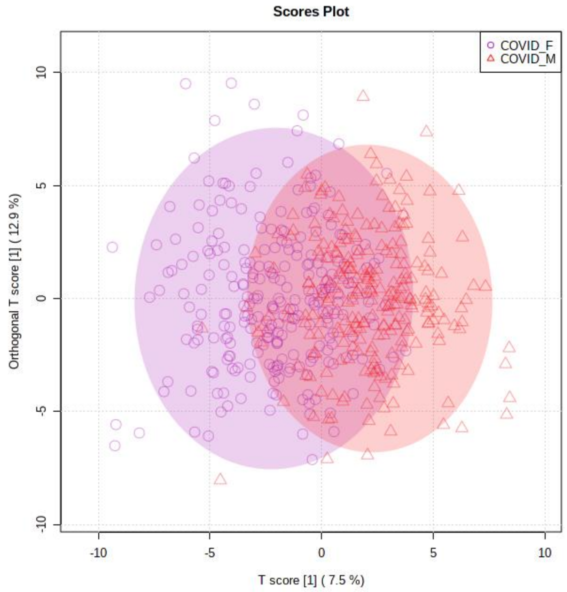


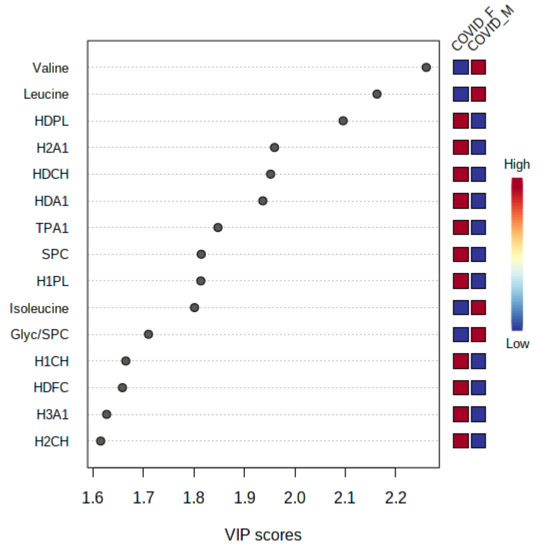


Analysis: COVID-19 + AHT: Female vs. Male

Univariate Analysis (sorted by p-value) (S8 Table)

| **Parameter** | **FC** | **log2(FC)** | **p.ajusted** | **-log10(p)** |
|---|---|---|---|---|
| Krea | 1.606 | 0.68347 | 4.85E-15 | 14.314 |
| Creatinine | 1.4297 | 0.51567 | 1.95E-11 | 10.711 |
| Leucine | 1.3474 | 0.43012 | 1.23E-08 | 7.9091 |
| Urea | 1.566 | 0.64708 | 2.61E-07 | 6.5835 |
| L1CH | 0.80279 | -0.31691 | 4.66E-05 | 4.3312 |
| L1PL | 0.81415 | -0.29664 | 4.70E-05 | 4.3276 |
| L1PN | 0.82365 | -0.27989 | 6.58E-05 | 4.1821 |
| L1AB | 0.82366 | -0.27988 | 6.58E-05 | 4.1821 |
| N-Dimethylglycine | 0.46741 | -1.0972 | 0.00019045 | 3.7202 |
| Citric acid | 1.2752 | 0.35067 | 0.00022161 | 3.6544 |
| L1FC | 0.81741 | -0.29087 | 0.00037996 | 3.4203 |
| L3CH | 0.76204 | -0.39206 | 0.0006598 | 3.1806 |
| Isoleucine | 1.2798 | 0.35595 | 0.00067522 | 3.1706 |
| L3AB | 0.78239 | -0.35404 | 0.0038371 | 2.416 |
| L3PN | 0.78511 | -0.34903 | 0.0039797 | 2.4002 |
| L4FC | 0.7936 | -0.33351 | 0.004768 | 2.3217 |
| L3PL | 0.77541 | -0.36697 | 0.0061644 | 2.2101 |
| L2PL | 0.82578 | -0.27618 | 0.0061644 | 2.2101 |
| L2AB | 0.83185 | -0.26561 | 0.0078533 | 2.1049 |
| L3FC | 0.79968 | -0.3225 | 0.0096501 | 2.0155 |
| Creatine | 0.55327 | -0.85395 | 0.012245 | 1.9121 |
| V2FC | 1.2274 | 0.29558 | 0.014255 | 1.846 |
| V3PL | 1.2118 | 0.27712 | 0.014255 | 1.846 |
| Dimethylsulfone | 1.2904 | 0.3678 | 0.020641 | 1.6853 |
| V2PL | 1.2261 | 0.29411 | 0.025487 | 1.5937 |

### Multivariate Analysis: COVID-19 + AHT: Female vs. Male (S9)

#### PCA

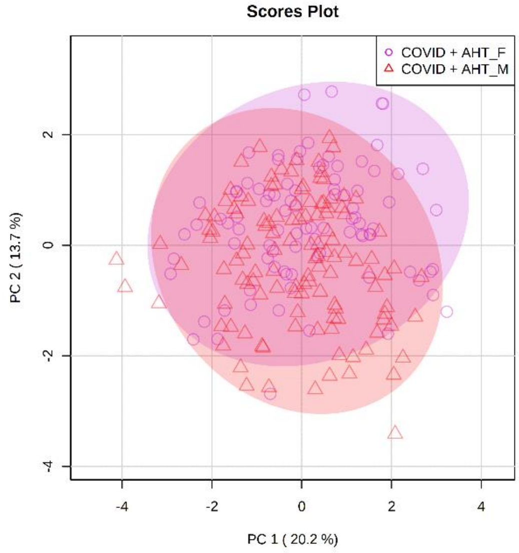


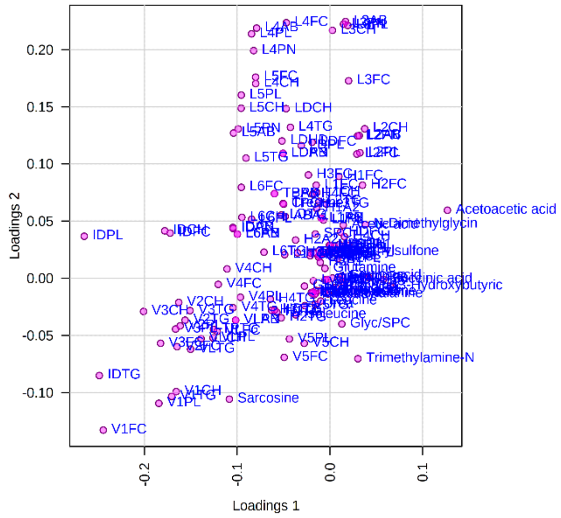


#### OPLS-DA

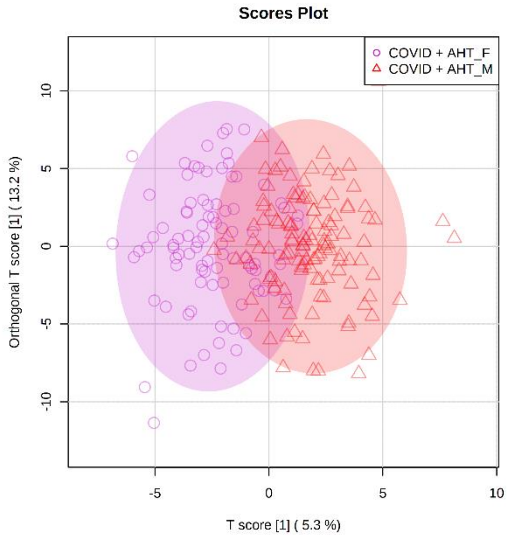


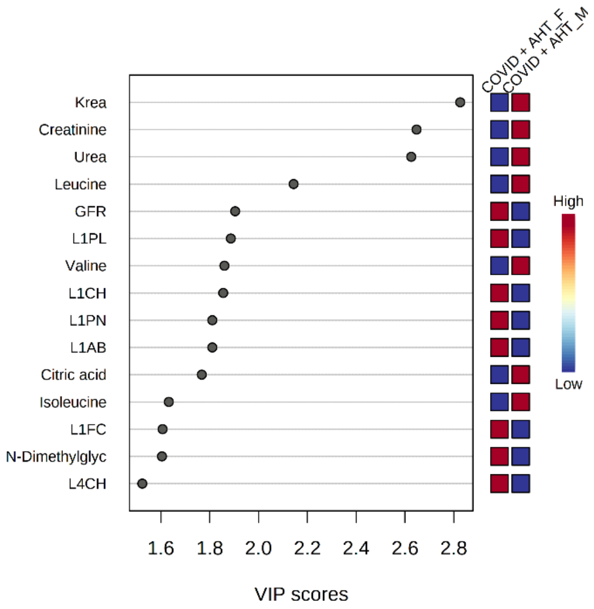

OPLS-DA (Creatinine, Kreatinin excluded)

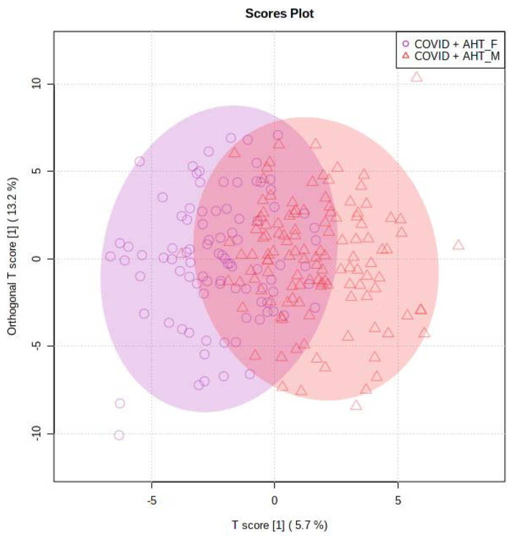


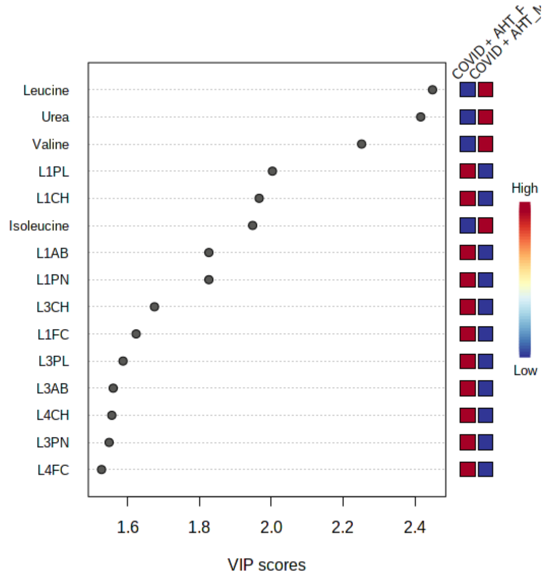


Analysis: Female: AHT vs. COVID-19 + AHT

Univariate Analysis (sorted by p-value) (S10 Table)

| Parameter | FC | log2(FC) | p.ajusted | -log10(p) |
|---|---|---|---|---|
| Pyruvic acid | 0.34299 | -1.5437 | 5.63E-17 | 16.249 |
| CRP | 4.3692 | 2.1274 | 6.47E-15 | 14.189 |
| Lysine | 0.62592 | -0.67594 | 5.55E-14 | 13.255 |
| Citric acid | 0.56397 | -0.82631 | 6.48E-11 | 10.189 |
| Glutamine | 0.78078 | -0.35701 | 3.68E-08 | 7.4343 |
| SPC | 0.73007 | -0.4539 | 5.42E-08 | 7.2658 |
| TPA1 | 0.78866 | -0.34253 | 5.42E-08 | 7.2658 |
| HDA1 | 0.79603 | -0.3291 | 2.46E-07 | 6.6098 |
| Leuk | 0.67381 | -0.56958 | 4.86E-07 | 6.313 |
| Alanine | 0.72843 | -0.45715 | 4.86E-07 | 6.313 |
| HDCH | 0.76998 | -0.37711 | 9.45E-07 | 6.0246 |
| TPA2 | 0.80433 | -0.31414 | 2.76E-06 | 5.5595 |
| Isoleucine | 0.64644 | -0.62942 | 5.11E-06 | 5.2915 |
| H4PL | 0.76104 | -0.39395 | 5.11E-06 | 5.2915 |
| HDPL | 0.80272 | -0.31703 | 5.11E-06 | 5.2915 |
| H4A1 | 0.77228 | -0.3728 | 5.52E-06 | 5.2578 |
| HDA2 | 0.82873 | -0.27102 | 1.11E-05 | 4.9558 |
| Glyc/SPC | 1.4629 | 0.54881 | 1.22E-05 | 4.9128 |
| Formic acid | 0.52923 | -0.91802 | 2.11E-05 | 4.6757 |
| H4CH | 0.72215 | -0.46964 | 4.90E-05 | 4.3095 |
| H2A1 | 0.80784 | -0.30786 | 8.04E-05 | 4.0949 |
| Leucine | 0.7479 | -0.41909 | 0.00014702 | 3.8326 |
| H3CH | 0.8059 | -0.31133 | 0.0001662 | 3.7794 |
| H4A2 | 0.75402 | -0.40733 | 0.0001669 | 3.7775 |
| TPCH | 0.81871 | -0.28858 | 0.00064526 | 3.1903 |
| H1CH | 0.72735 | -0.45928 | 0.0008657 | 3.0626 |
| L6PL | 0.78795 | -0.34382 | 0.00231 | 2.6364 |

| | | | | |
|---|---|---|---|---|
| Dimethylsulfone | 0.6156 | -0.69993 | 0.0025857 | 2.5874 |
| L6CH | 0.76671 | -0.38325 | 0.0027105 | 2.5669 |
| H1FC | 0.7069 | -0.50042 | 0.002792 | 2.5541 |
| L6FC | 0.74434 | -0.42597 | 0.0031034 | 2.5082 |
| H3FC | 0.72268 | -0.46857 | 0.0034169 | 2.4664 |
| IDPL | 0.64342 | -0.63616 | 0.0069898 | 2.1555 |
| Tyrosine | 0.74021 | -0.43399 | 0.0076518 | 2.1162 |
| HDFC | 0.80722 | -0.30896 | 0.009037 | 2.044 |
| L6AB | 0.80154 | -0.31916 | 0.0093814 | 2.0277 |
| L6PN | 0.80159 | -0.31907 | 0.0093814 | 2.0277 |
| 3-Hydroxybutyric acid | 0.47148 | -1.0847 | 0.012008 | 1.9205 |
| H1PL | 0.77134 | -0.37456 | 0.012392 | 1.9068 |
| Glutamic acid | 1.567 | 0.64805 | 0.014425 | 1.8409 |
| Sarcosine | 0.64083 | -0.642 | 0.014882 | 1.8273 |
| L5CH | 0.74147 | -0.43155 | 0.019358 | 1.7131 |
| IDTG | 0.66494 | -0.5887 | 0.021925 | 1.6591 |
| H1A1 | 0.76831 | -0.38024 | 0.027082 | 1.5673 |
| H4FC | 0.76722 | -0.38229 | 0.03287 | 1.4832 |
| L5PL | 0.78218 | -0.35443 | 0.039143 | 1.4073 |

Multivariate Analysis: AHT vs. COVID-19 + AHT: Female (S11)

PCA

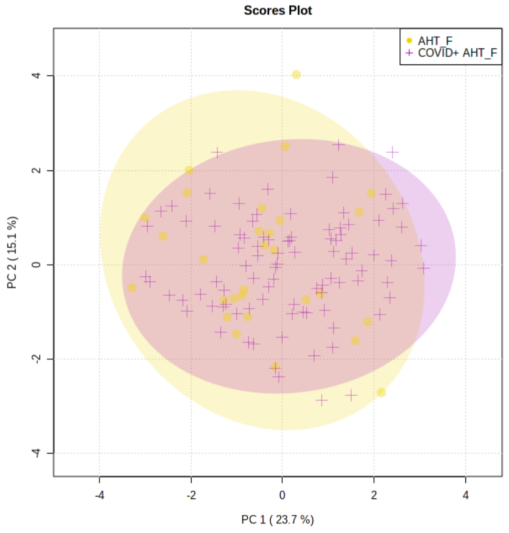


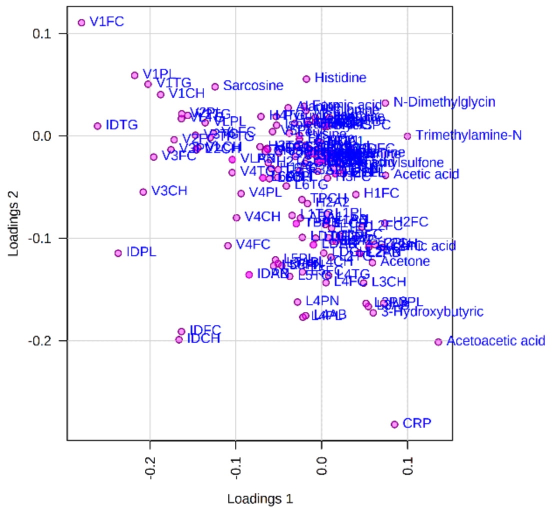

OPLS-DA

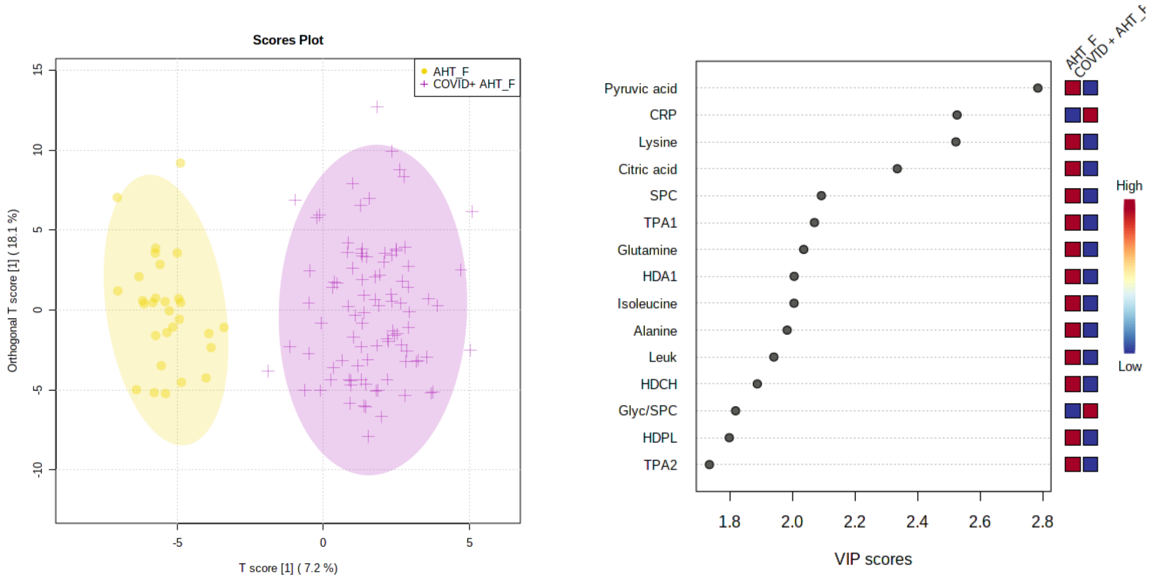


Analysis: Male: AHT vs. COVID-19 + AHT

Univariate Analysis (sorted by p-value) (S12 Table)

| Parameter | FC | log2(FC) | p.ajusted | -log10(p) |
|---|---|---|---|---|
| Pyruvic acid | 0.34485 | -1.536 | 3.52E-19 | 18.454 |
| CRP | 4.2723 | 2.095 | 1.57E-16 | 15.803 |
| Lysine | 0.63734 | -0.64987 | 8.95E-11 | 10.048 |
| Glutamine | 0.72924 | -0.45554 | 3.94E-09 | 8.4046 |
| H4A1 | 0.75654 | -0.40252 | 6.00E-08 | 7.2221 |
| H4PL | 0.75084 | -0.41343 | 1.08E-07 | 6.9663 |
| Alanine | 0.76222 | -0.39173 | 2.04E-07 | 6.6896 |
| H4A2 | 0.71879 | -0.47635 | 2.17E-07 | 6.6644 |
| Formic acid | 0.4412 | -1.1805 | 2.77E-07 | 6.557 |
| Isoleucine | 0.65948 | -0.6006 | 5.23E-06 | 5.2817 |
| TPCH | 0.73219 | -0.44971 | 5.23E-06 | 5.2817 |
| Methionine | 0.72368 | -0.46658 | 1.23E-05 | 4.9085 |
| H4CH | 0.71043 | -0.49324 | 8.90E-05 | 4.0504 |
| Glyc/SPC | 1.4649 | 0.55084 | 0.00012528 | 3.9021 |
| Leuk | 0.71046 | -0.49318 | 0.00026453 | 3.5775 |
| SPC | 0.81147 | -0.3014 | 0.0002678 | 3.5722 |
| L5CH | 0.5579 | -0.84191 | 0.00060334 | 3.2194 |
| L6CH | 0.6593 | -0.60099 | 0.00060334 | 3.2194 |
| IDPL | 0.52262 | -0.93616 | 0.00073102 | 3.1361 |
| L2TG | 1.284 | 0.36068 | 0.00083945 | 3.076 |
| L6PN | 0.66376 | -0.59128 | 0.0011325 | 2.946 |
| L6AB | 0.66839 | -0.58125 | 0.0013503 | 2.8696 |
| L5AB | 0.5913 | -0.75803 | 0.0013572 | 2.8674 |
| L5PN | 0.61138 | -0.70985 | 0.0015575 | 2.8076 |
| L5PL | 0.59748 | -0.74303 | 0.0016275 | 2.7885 |

| | | | | |
|---|---|---|---|---|
| L6PL | 0.67226 | -0.5729 | 0.0016275 | 2.7885 |
| Leucine | 0.79928 | -0.32324 | 0.0016275 | 2.7885 |
| V1FC | 0.54109 | -0.88605 | 0.0023185 | 2.6348 |
| LDPL | 0.74792 | -0.41905 | 0.0023185 | 2.6348 |
| Histidine | 0.71859 | -0.47676 | 0.0026369 | 2.5789 |
| TPAB | 0.78371 | -0.35161 | 0.0026369 | 2.5789 |
| TBPN | 0.78371 | -0.35161 | 0.0026369 | 2.5789 |
| L6FC | 0.64845 | -0.62494 | 0.0034598 | 2.4609 |
| Citric acid | 0.80975 | -0.30445 | 0.0037056 | 2.4311 |
| V1TG | 0.61227 | -0.70777 | 0.0038169 | 2.4183 |
| Glutamic acid | 1.4374 | 0.5235 | 0.0038772 | 2.4115 |
| LDCH | 0.69071 | -0.53385 | 0.0042782 | 2.3687 |
| LDPN | 0.75427 | -0.40685 | 0.0042782 | 2.3687 |
| LDAB | 0.75427 | -0.40685 | 0.0042782 | 2.3687 |
| V1PL | 0.58875 | -0.76427 | 0.0047406 | 2.3242 |
| L4AB | 0.60458 | -0.72599 | 0.0047406 | 2.3242 |
| L5FC | 0.6296 | -0.66749 | 0.0047406 | 2.3242 |
| VLPL | 0.72496 | -0.46403 | 0.0050067 | 2.3004 |
| L4CH | 0.56466 | -0.82454 | 0.0055891 | 2.2527 |
| L4PN | 0.61193 | -0.70856 | 0.0055891 | 2.2527 |
| IDFC | 0.66339 | -0.59207 | 0.0055891 | 2.2527 |
| VLTG | 0.67483 | -0.5674 | 0.0099364 | 2.0028 |
| L1CH | 0.781 | -0.35661 | 0.010783 | 1.9673 |
| H2TG | 1.2388 | 0.30893 | 0.011086 | 1.9552 |
| IDTG | 0.51742 | -0.95058 | 0.011198 | 1.9509 |
| V1CH | 0.65974 | -0.60003 | 0.011198 | 1.9509 |
| L4FC | 0.64492 | -0.6328 | 0.011887 | 1.9249 |
| Sarcosine | 0.56744 | -0.81747 | 0.012715 | 1.8957 |
| Tyrosine | 0.82192 | -0.28293 | 0.013507 | 1.8694 |
| IDCH | 0.66296 | -0.593 | 0.015684 | 1.8046 |
| VLFC | 0.76758 | -0.38161 | 0.015684 | 1.8046 |
| H4FC | 0.76955 | -0.37791 | 0.015684 | 1.8046 |
| LDFC | 0.77818 | -0.36183 | 0.015684 | 1.8046 |
| L4PL | 0.59681 | -0.74465 | 0.017443 | 1.7584 |
| TPTG | 0.70496 | -0.50438 | 0.017822 | 1.749 |
| Succinic acid | 2.7587 | 1.464 | 0.018408 | 1.735 |
| L1FC | 0.80636 | -0.31051 | 0.024057 | 1.6188 |
| Krea | 1.3017 | 0.38036 | 0.026281 | 1.5804 |
| Lactic acid | 1.2756 | 0.35117 | 0.026326 | 1.5796 |
| H4TG | 0.81799 | -0.28984 | 0.049361 | 1.3066 |

### Multivariate Analysis: AHT vs. COVID-19 + AHT: Male (S13)

#### PCA

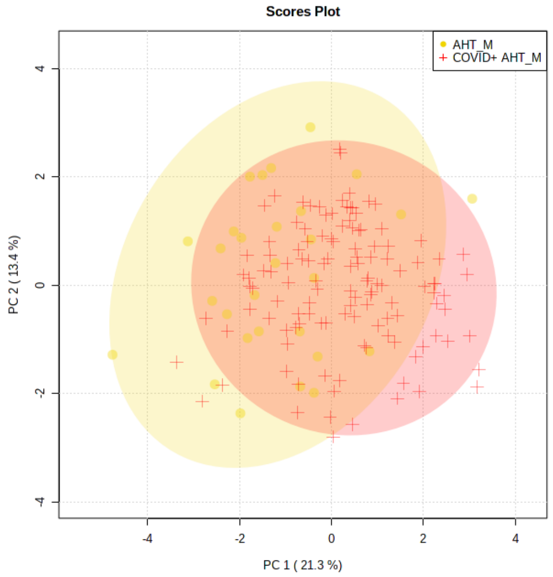


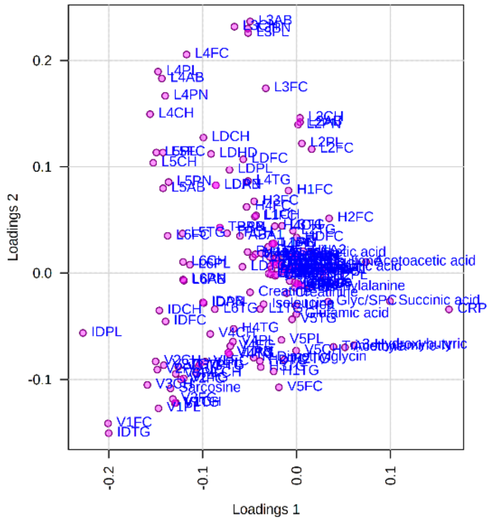


#### OPLS-DA

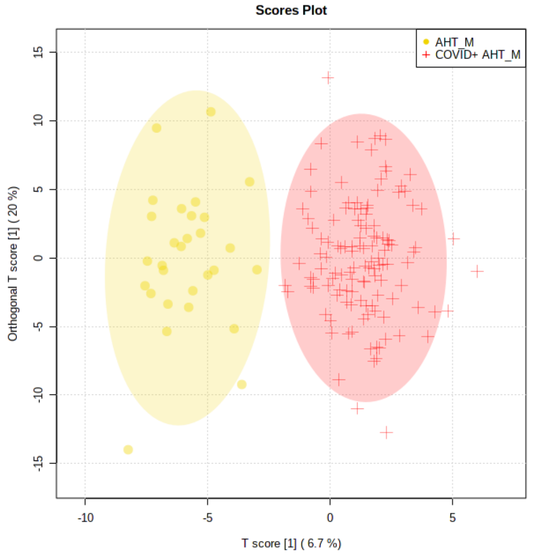


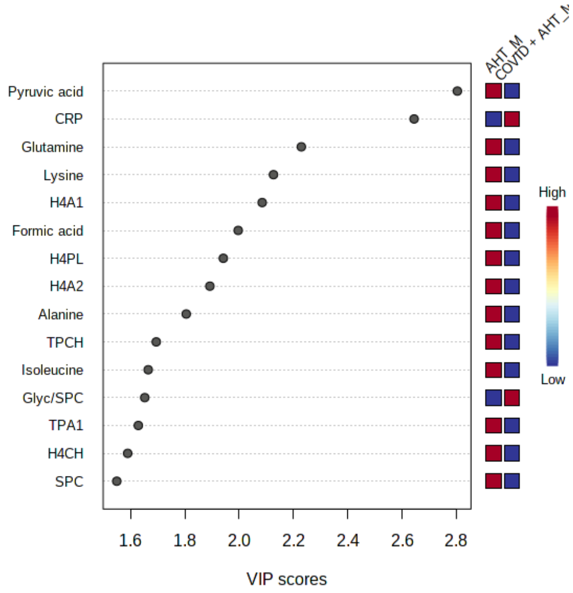


### Analysis: COVID-19: AHT vs. regular BP

#### Univariate Analysis (sorted by p-value) (S14 Table)

| Parameter | FC | log2(FC) | p.ajusted | -log10(p) |
|---|---|---|---|---|
| Urea | 1.3859 | 0.47079 | 2.31E-10 | 9.6361 |
| Creatinine | 1.2227 | 0.29007 | 8.97E-08 | 7.0474 |
| Krea | 1.2426 | 0.31336 | 8.93E-07 | 6.0489 |
| TPTG | 1.2247 | 0.29246 | 9.98E-05 | 4.0008 |
| L3PL | 0.818 | -0.28982 | 0.00013043 | 3.8846 |

| | | | | |
|---|---|---|---|---|
| VLTG | 1.2554 | 0.3281 | 0.00020075 | 3.6974 |
| IDTG | 1.4217 | 0.50758 | 0.00023391 | 3.6309 |
| V1TG | 1.3416 | 0.42396 | 0.00028581 | 3.5439 |
| V1PL | 1.3418 | 0.42418 | 0.00029831 | 3.5253 |
| V1CH | 1.3136 | 0.39355 | 0.000543 | 3.2652 |
| L3CH | 0.78596 | -0.34747 | 0.00056694 | 3.2465 |
| V2TG | 1.229 | 0.29748 | 0.0019191 | 2.7169 |
| Dimethylsulfone | 0.51527 | -0.9566 | 0.0032443 | 2.4889 |
| V2FC | 1.2307 | 0.29945 | 0.0032443 | 2.4889 |
| V3FC | 1.2446 | 0.3157 | 0.003337 | 2.4766 |
| V2PL | 1.2106 | 0.27577 | 0.003337 | 2.4766 |
| Trimethylamine-N-oxide | 1.2593 | 0.33267 | 0.012914 | 1.8889 |
| Succinic acid | 1.4183 | 0.50414 | 0.038851 | 1.4106 |

Multivariate Analysis: COVID-19: AHT vs. regular BP (S15)

PCA

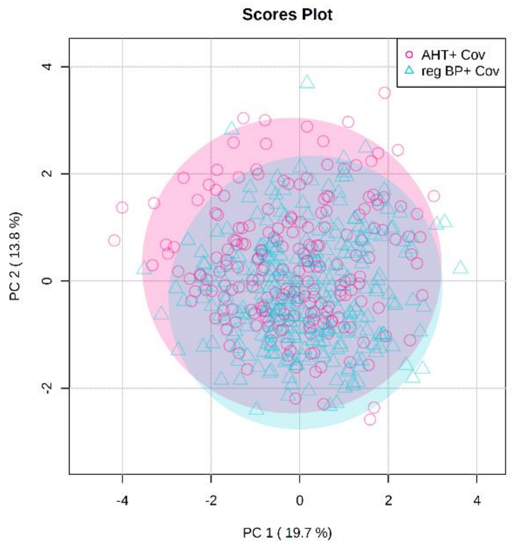


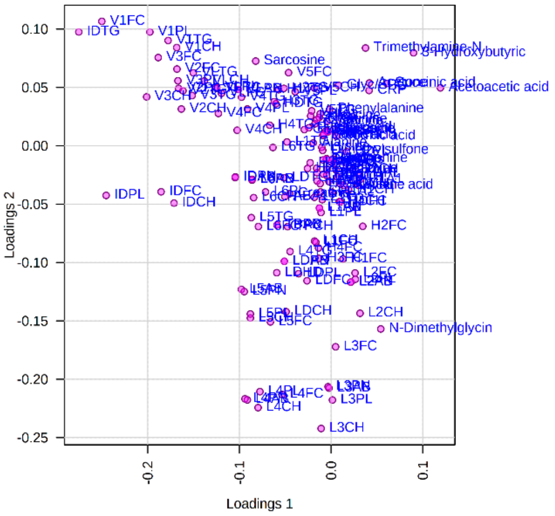

OPLS-DA

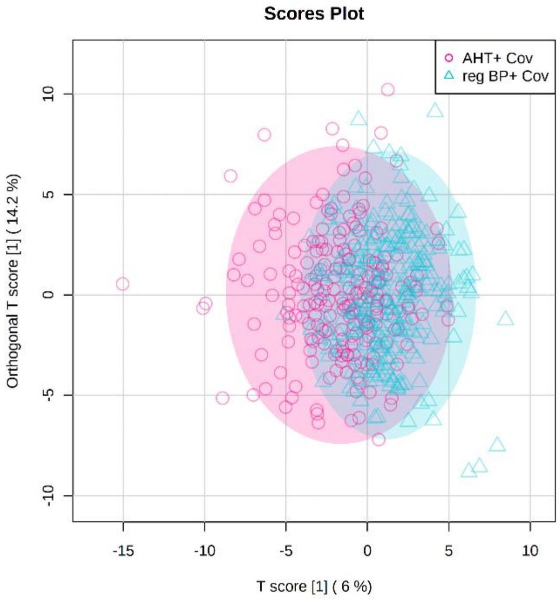


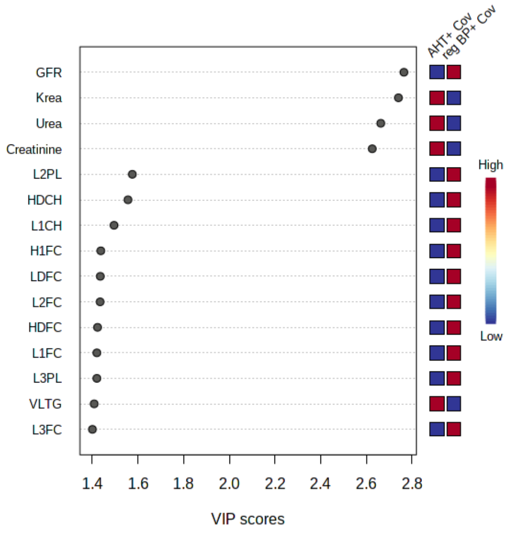


Analysis: Hospitalized COVID-19 patients: AHT vs. regular BP

Univariate Analysis (sorted by p-value) (S16 Table)

| Parameter | FC | log2(FC) | p.ajusted | -log10(p) |
|---|---|---|---|---|
| Krea | 1.5054 | 0.59016 | 0.00047905 | 3.3196 |
| Creatinine | 1.3493 | 0.43221 | 0.00047905 | 3.3196 |
| Urea | 1.6369 | 0.71095 | 0.0082981 | 2.081 |
| Creatine | 0.49471 | -1.0153 | 0.012673 | 1.8971 |
| V1CH | 1.478 | 0.5636 | 0.012673 | 1.8971 |
| V1TG | 1.4507 | 0.5368 | 0.012673 | 1.8971 |
| H2TG | 1.263 | 0.33684 | 0.012673 | 1.8971 |
| HDTG | 1.2326 | 0.30176 | 0.012673 | 1.8971 |
| H3TG | 1.2303 | 0.29896 | 0.012673 | 1.8971 |
| V2FC | 1.4265 | 0.51248 | 0.020486 | 1.6885 |
| VLTG | 1.3405 | 0.42273 | 0.020486 | 1.6885 |
| TPTG | 1.3085 | 0.3879 | 0.020486 | 1.6885 |
| H1TG | 1.2993 | 0.3777 | 0.020486 | 1.6885 |
| VLAB | 1.2303 | 0.29905 | 0.020486 | 1.6885 |
| VLPN | 1.2303 | 0.299 | 0.020486 | 1.6885 |
| V1PL | 1.4318 | 0.51783 | 0.023103 | 1.6363 |
| V3FC | 1.4485 | 0.53451 | 0.029656 | 1.5279 |
| IDTG | 1.6412 | 0.71478 | 0.04039 | 1.3937 |

Multivariate Analysis: Hospitalized COVID-19 patients: AHT vs. regular BP (S17)

PCA

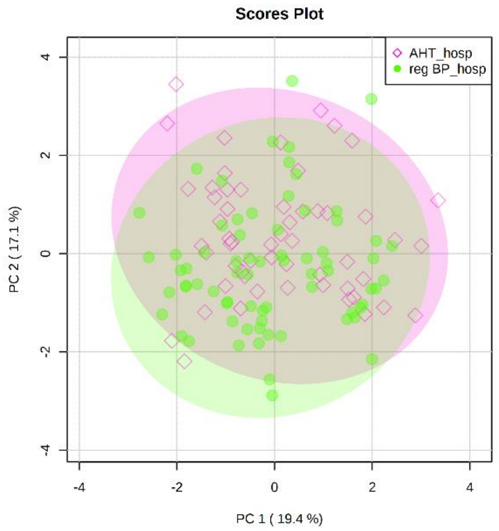


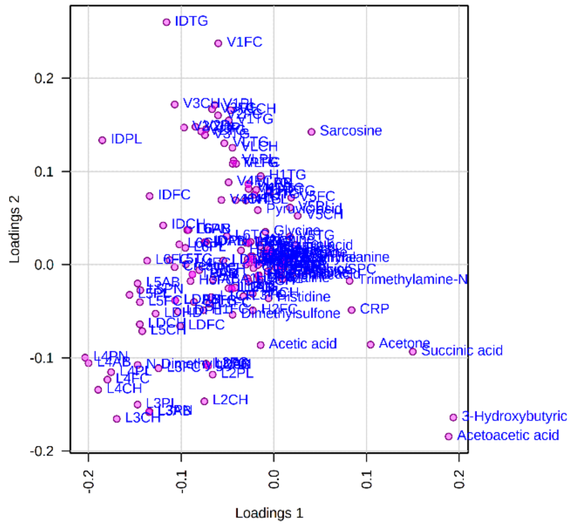


OPLS-DA

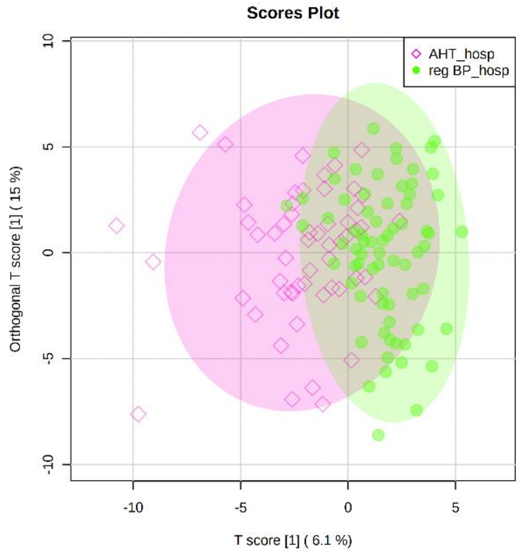


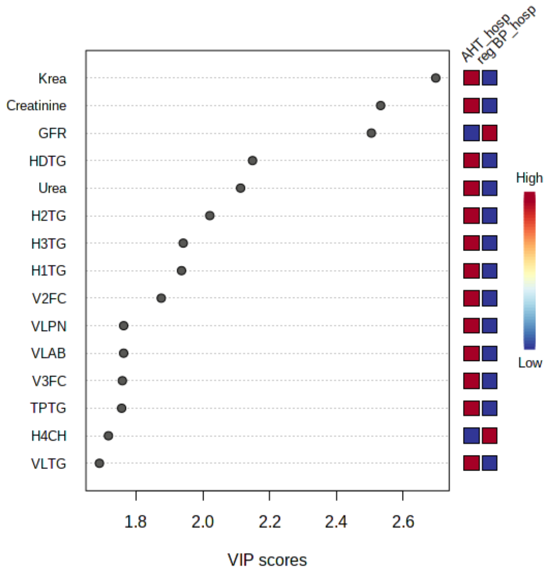


Analysis: COVID-19 + AHT: RAASI vs. noRAASI

Univariate Analysis (sorted by p-value) (S18 Table)

| Parameter | FC | log2(FC) | p.ajusted | -log10(p) |
|---|---|---|---|---|
| Creatinine | 1.4132 | 0.499 | 0.012372 | 1.9075 |
| GFR | 0.82702 | -0.27401 | 0.012372 | 1.9075 |
| Pyruvic acid | 1.3973 | 0.48267 | 0.014568 | 1.8366 |

| | | | | |
|---|---|---|---|---|
| V4TG | 1.4137 | 0.49952 | 0.019904 | 1.7011 |
| V4PL | 1.3766 | 0.46108 | 0.022486 | 1.6481 |
| VLPN | 1.3316 | 0.41317 | 0.026222 | 1.5813 |
| VLAB | 1.3315 | 0.41309 | 0.026222 | 1.5813 |
| Krea | 1.5653 | 0.64644 | 0.026385 | 1.5786 |
| V4CH | 1.435 | 0.52105 | 0.039837 | 1.3997 |

Multivariate Analysis: COVID-19 + AHT: RAASI vs. noRAASI (S19)

PCA

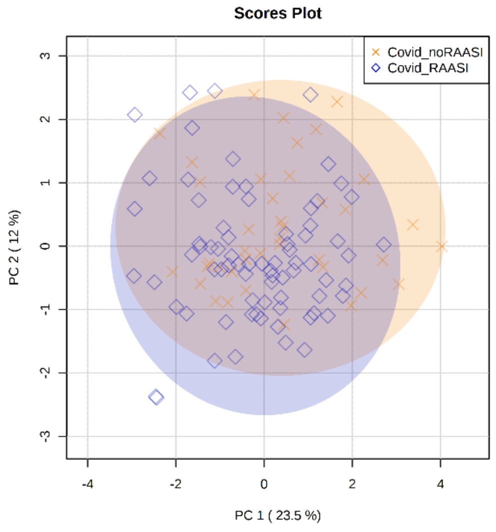


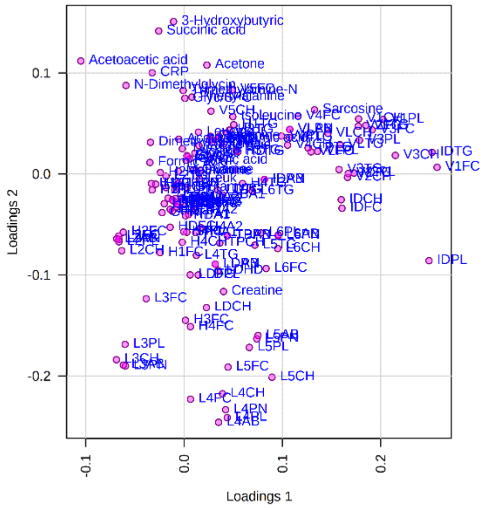


OPLS-DA

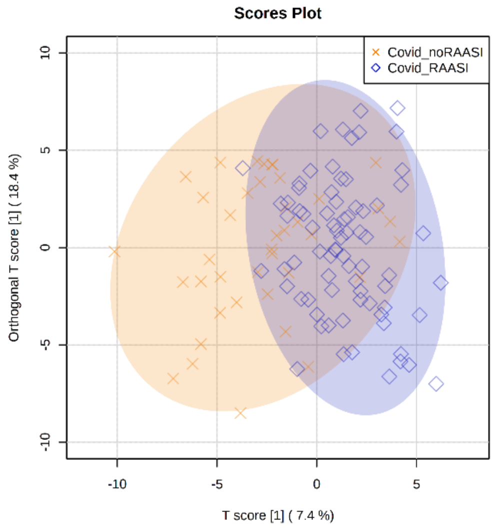


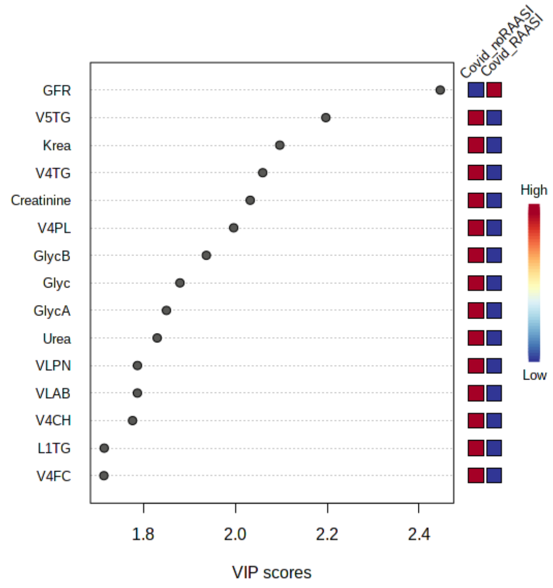

Violin plots and unpaired t-tests, showing typical altered metabolites and lipoproteins in COVID-19 patients (S20)

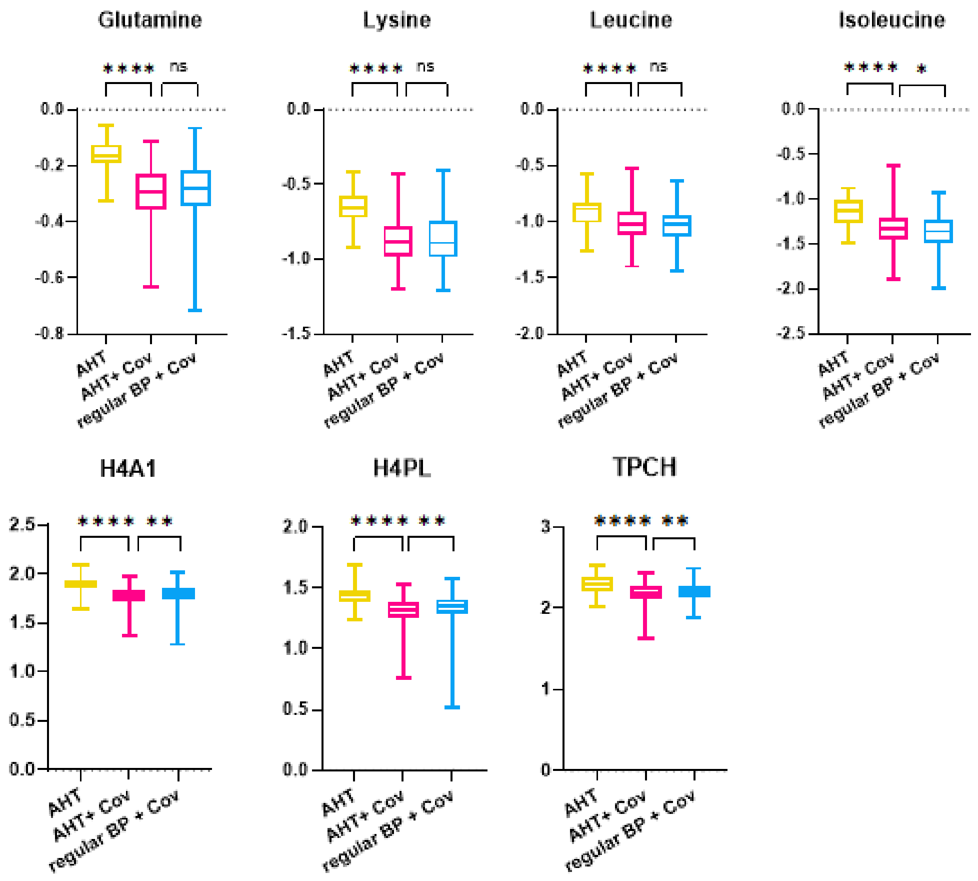


### Abbreviations (S21 Table)

| **Name** | **Extended name** |
|---|---|
| ACEI | Angiotensin- converting- enzyme inhibitor |
| AHT | Arterial hypertension |
| AT1RB | Angiotensin-1 receptor blockers |
| AUC | Area under the curve |
| BB | Beta blockers |
| BCAA | Branched chained amino acids (leucine, isoleucine, valine) |
| BP | Blood pressure |
| CCB | Calcium channel blockers |
| FC | Fold change |
| FDR | False Discovery rate |
| HC | Healthy cohort |
| NMR | Nuclear magnetic resonance |
| T2DM | Diabetes mellitus Type 2 |
| PACS | Post-acute-covid- syndrom |
| PCA | Principal component analysis |
| RAASI | Renin-angiotensin-aldosterone- system inhibitors |

| Name | Extended name | Unit |
|---|---|---|
| BUN | Blood urea nitrogen | mg/dL |
| Creatinine.lab | Creatinine (laboratory report) | mg/dL |
| CRP | C- reactive protein | mg/L |
| GFR | Glomerular filtration rate | ml/min |
| GGT | Gamma-glutamyl transferase | Unit/L |
| Glyc | Glycoproteins | Procedure defined Unit |
| GOT | Glutamate- oxalacetate- transferase | Unit/L |
| Leuk | Leucozytes | $x10^9$cells/L |
| LDH | Lactat dehydrogenase | Unit/L |
| SPC | Supramolecular phospholipid composite | Procedure defined Unit |
| TMAO | Trimethylamine-N-oxide | mmol/L |

| Name | Extended name | Unit |
|---|---|---|
| ABA1 | Apolipoprotein- B100/Apolipoprotein-A1 | none |
| H1A1 | Apolipoprotein- A1 HDL-1 | mg/dL |
| H1A2 | Apolipoprotein- A2 HDL-1 | mg/dL |
| H1CH | Cholesterol HDL-1 | mg/dL |
| H1FC | Free Cholesterol HDL-1 | mg/dL |
| H1PL | Phospholipids HDL-1 | mg/dL |
| H1TG | Triglycerides HDL-1 | mg/dL |
| H2A1 | Apolipoprotein- A1 HDL-2 | mg/dL |
| H2A2 | Apolipoprotein- A2 HDL-2 | mg/dL |
| H2CH | Cholesterol HDL-2 | mg/dL |
| H2FC | Free Cholesterol HDL-2 | mg/dL |
| H2PL | Phospholipids HDL-2 | mg/dL |
| H2TG | Triglycerides HDL-2 | mg/dL |
| H3A1 | Apolipoprotein- A1 HDL-3 | mg/dL |
| H3A2 | Apolipoprotein- A2 HDL-3 | mg/dL |
| H3CH | Cholesterol HDL-3 | mg/dL |
| H3FC | Free Cholesterol HDL-3 | mg/dL |
| H3PL | Phospholipids HDL-3 | mg/dL |
| H3TG | Triglycerides HDL-3 | mg/dL |
| H4A1 | Apolipoprotein- A1 HDL-4 | mg/dL |
| H4A2 | Apolipoprotein- A2 HDL-4 | mg/dL |
| H4CH | Cholesterol HDL-4 | mg/dL |
| H4FC | Free Cholesterol HDL-4 | mg/dL |
| H4PL | Phospholipids HDL-4 | mg/dL |
| H4TG | Triglycerides HDL-4 | mg/dL |
| HDA1 | HDL-Apolipoprotein-A1 | mg/dL |
| HDA2 | HDL-Apolipoprotein-A2 | mg/dL |
| HDCH | HDL-Cholesterol | mg/dL |
| HDFC | HDL Free Cholesterol | mg/dL |
| HDPL | HDL Phospholipids | mg/dL |
| HDTG | HDL Triglycerides | mg/dL |
| IDAB | IDL-Apolipoprotein-B100 | mg/dL |
| IDCH | IDL Cholesterol | mg/dL |
| IDFC | IDL Free Cholesterol | mg/dL |
| IDPL | IDL Phospholipids | mg/dL |
| IDPN | IDL Particle Numer | mg/dL |

| | | |
|---|---|---|
| IDTG | IDL Triglycerides | mg/dL |
| L1AB | Apolipoprotein-B100 LDL-1 | mg/dL |
| L1CH | Cholesterol LDL-1 | mg/dL |
| L1FC | Free Cholesterol LDL-1 | mg/dL |
| L1PL | Phospholipids LDL-1 | mg/dL |
| L1PN | Particle Number LDL-1 | mg/dL |
| L1TG | Triglycerides LDL-1 | mg/dL |
| L2AB | Apolipoprotein-B100 LDL-2 | mg/dL |
| L2CH | Cholesterol LDL-2 | mg/dL |
| L2FC | Free Cholesterol LDL-2 | mg/dL |
| L2PL | Phospholipids LDL-2 | mg/dL |
| L2PN | Particle Number LDL-2 | mg/dL |
| L2TG | Triglycerides LDL-2 | mg/dL |
| L3AB | Apolipoprotein-B100 LDL-3 | mg/dL |
| L3CH | Cholesterol LDL-3 | mg/dL |
| L3FC | Free Cholesterol LDL-3 | mg/dL |
| L3PL | Phospholipids LDL-3 | mg/dL |
| L3PN | Particle Number LDL-3 | mg/dL |
| L3TG | Triglycerides LDL-3 | mg/dL |
| L4AB | Apolipoprotein-B100 LDL-4 | mg/dL |
| L4CH | Cholesterol LDL-4 | mg/dL |
| L4FC | Free Cholesterol LDL-4 | mg/dL |
| L4PL | Phospholipids LDL-4 | mg/dL |
| L4PN | Particle Number LDL-4 | mg/dL |
| L4TG | Triglycerides LDL-4 | mg/dL |
| L5AB | Apolipoprotein-B100 LDL-5 | mg/dL |
| L5CH | Cholesterol LDL-5 | mg/dL |
| L5FC | Free Cholesterol LDL-5 | mg/dL |
| L5PL | Phospholipids LDL-5 | mg/dL |
| L5PN | Particle Number LDL-5 | mg/dL |
| L5TG | Triglycerides LDL-5 | mg/dL |
| L6AB | Apolipoprotein-B100 LDL-6 | mg/dL |
| L6CH | Cholesterol LDL-6 | mg/dL |
| L6FC | Free Cholesterol LDL-6 | mg/dL |
| L6PL | Phospholipids LDL-6 | mg/dL |
| L6PN | Particle Number LDL-6 | mg/dL |
| L6TG | Triglycerides LDL-6 | mg/dL |
| LDAB | LDL- Apolipoprotein-B100 | mg/dL |
| LDCH | LDL Cholesterol | mg/dL |
| LDFC | LDL Free Cholesterol | mg/dL |
| LDHD | LDL Cholesterol/HDL-Cholesterol | mg/dL |
| LDPL | LDL Phospholipids | mg/dL |
| LDPN | LDL Particle Number | mg/dL |
| LDTG | LDL Triglycerides | mg/dL |
| TBPN | Total Particle Numer (apolipoprotein-B100 carrying particles) | mg/dL |
| TPA1 | Total Plasma Apolipoprotein-A1 | mg/dL |
| TPA2 | Total Plasma Apolipoprotein-A2 | mg/dL |
| TPAB | Total Plasma Apolipoprotein-B100 | mg/dL |
| TPCH | Total Plasma Cholesterol | mg/dL |
| TPTG | Total Plasma Triglycerides | mg/dL |

| | | |
|---|---|---|
| V1CH | Cholesterol VLDL-1 | mg/dL |
| V1FC | Free Cholesterol VLDL-1 | mg/dL |
| V1PL | Phospholipids VLDL-1 | mg/dL |
| V1TG | Triglycerides VLDL-1 | mg/dL |
| V2CH | Cholesterol VLDL-2 | mg/dL |
| V2FC | Free Cholesterol VLDL-2 | mg/dL |
| V2PL | Phospholipids VLDL-2 | mg/dL |
| V2TG | Triglycerides VLDL-2 | mg/dL |
| V3CH | Cholesterol VLDL-3 | mg/dL |
| V3FC | Free Cholesterol VLDL-3 | mg/dL |
| V3PL | Phospholipids VLDL-3 | mg/dL |
| V3TG | Triglycerides VLDL-3 | mg/dL |
| V4CH | Cholesterol VLDL-4 | mg/dL |
| V4FC | Free Cholesterol VLDL-4 | mg/dL |
| V4PL | Phospholipids VLDL-4 | mg/dL |
| V4TG | Triglycerides VLDL-4 | mg/dL |
| V5CH | Cholesterol VLDL-5 | mg/dL |
| V5FC | Free Cholesterol VLDL-5 | mg/dL |
| V5PL | Phospholipids VLDL-5 | mg/dL |
| V5TG | Triglycerides VLDL-5 | mg/dL |
| VLAB | VLDL-Apolipoprotein-B100 | mg/dL |
| VLCH | VLDL Cholesterol | mg/dL |
| VLFC | VLDL Free Cholesterol | mg/dL |
| VLPL | VLDL Phospholipids | mg/dL |
| VLPN | VLDL Particle Number | mg/dL |
| VLTG | VLDL Triglycerides | mg/dL |
